# Phylogenetic study of local patterns influenza A(H3N2) virus transmission in a semi-isolated population in a remote island in Japan between 2011-2013

**DOI:** 10.1101/2024.05.14.24307330

**Authors:** Su Myat Han, Teiichiro Shiino, Shingo Masuda, Yuki Furuse, Takahiro Yasaka, Satoshi Kanda, Kazuhiri Komori, Nobuo Saito, Yoshiano Kubo, Chris Smith, Akira Endo, Alexis Robert, Marc Baguelin, Koya Ariyoshi

## Abstract

**Background:** The spatial spread and importation risk of influenza A viruses in rural settings remains unclear due to the sparsity of representative spatiotemporal sequence data.

**Methods:** Nasopharyngeal (NPS) samples of Rapid Influenza Diagnostic Test (RIDT) positive individuals in Kamigoto Island, Japan, were confirmed using quantitative polymerase chain reaction (RT-PCR). The confirmed influenza A positive samples were processed for whole-genome sequencing. Time-resolved phylogenetic trees were built from HA sequences to classify the circulating clades, with events of introductions and local clustering. Spatio-temporal transmission patterns were then analyzed for the largest local clusters.

**Results:** We obtained 178 whole-genome sequences of influenza A/H3N2 collected during 2011/12 and 2012/13 influenza seasons. The time-resolved phylogenetic tree identified at least six independent introduction events in 2011/12 and 2012/13. Majority of Kamigoto strains are closely related to strains from mainland Japan. All 2011/12 strains were identified as clade 3 C.2 (n=29), while 2012/13 strains fell into two clades: clade 3C.2 (n=129), and 3C.3a (n=20). No local persistence over one year was observed for Kamigoto strains. The spatio-temporal analysis of the largest cluster revealed that the first case and a large number of cases came from the busiest district of the island and spread towards the other parts of the island.

**Conclusion:** Influenza A(H3N2) virus outbreaks in Kamigoto island were marked by multiple introductions and fueled by local transmission. All the identified clusters in 2012/13 season circulate simultaneously. These cases may be misinterpreted as part of the same cluster without sequencing data, highlighting the importance of genomic surveillance. The results of this study are based on a two-year analysis of influenza sequences from the island; repeated analyzes for different influenza seasons and geographic locations will help us better understand detailed transmission patterns.

## Introduction

Seasonal influenza remains a major public health threat, causing substantial morbidity and mortality annually. It is estimated that influenza affects 3-5 million people globally, resulting in approximately 290,000-650,000 deaths each year.^1^ The transmission patterns of seasonal influenza viruses have been extensively studied using surveillance data,^2–6^ which helped inform influenza seasonal outbreak dynamics, the type of strain circulating at each season, and vaccine selection based on genomic surveillance. It has also provided guidance on the optimal timing for vaccination.^3,7,8^

In Japan, the estimated annual prevalence of seasonal influenza exceeds 10% of the population.^9^ Japan is home to many smaller islands, some of which have limited connections to mainland Japan.^10^ Kamigoto Island is one of these, offering unique insights into localized transmission dynamics in an isolated setting. In a previous study conducted in Kamigoto island,^2^ we used influenza surveillance data containing cases confirmed via rapid influenza diagnostic test (RIDT) between 2010 and 2018 to identify determinants of transmission and local dynamics of influenza transmission. The study revealed patterns of transmission largely influenced by age, vaccine coverage, and district population density. However, surveillance data would not be able to distinguish between transmission chains co-circulating at the same time and areas, rendering accurate importation risk assessment challenging.

Given the diverse strains and subtypes within Influenza A that can cocirculate, reliance solely on influenza-like illness (ILI) and rapid influenza diagnostic test (RIDT) data often falls short in identifying distinct influenza strains. While a subset of samples from the surveillance data were processed for subtype identification (for example H3 or H1), this analysis alone cannot pinpoint the time and location of the virus’s introduction in the local population. Genomic sequence data can bridge this knowledge gap by offering a deeper understanding of influenza’s antigenic variability and their connections to various parts of the world.

Phylogenetic analysis is instrumental in reconstructing the evolutionary relationship between genomic sequences of infected individuals sampled at different dates, ^11–13^ and enables the identification of the epidemiological relationship between cases. Cases with similar genomic sequences are more likely to be epidemiologically connected, and different strains indicate separate importations. The time-resolved phylogeny can also be used to infer who- infected-whom among the sampled cases, thereby exploring the pathogen’s transmission dynamics.^11,12,14–16^ In recent years, the growing accessibility of sequence data has prompted extensive analysis during outbreaks to examining their origins and dynamics. Integrating genetic sequences with epidemiological contact tracing data has enriched outbreak investigations, yielding a better understanding of infection patterns when cases are densely sampled. However, in the case of influenza, only a small fraction of the whole samples is collected for further analysis during the influenza season, limiting its usefulness in reconstructing transmission chains.

This study used the nasopharyngeal swab (NPS) samples collected from influenza-like illness (ILI) patients presenting to the Kamigoto hospital during the 2011/12 and 2012/13 influenza seasons. We performed whole genome sequencing on reverse transcription-polymerase chain reaction (RT-PCR) confirmed influenza cases. We used these sequences and the date of collection of the cases to conduct phylogenetic analysis and understand the relationship of Kamigoto strains to those in mainland Japan and the rest of the world. Then, we assessed whether all cases were grouped in the same cluster or if they were related to independent importations and analyzed the role of importation versus local transmission within the island. Finally, we explored the temporal and spatial distribution of the clusters identified from the phylogenetic study.

## Methods

### Study design and setting

The samples used in this study were collected from patients recorded with ILI visiting to the Kamigoto hospital in Kamigoto island, Japan, during the 2011/12 and 2012/13 influenza seasons. Kamigoto Island is located on the western coast of Japan, with a population of 22,599 inhabitants in 2011.^17^ Kamigoto Hospital is the only hospital in the island with all levels of care (primary, secondary, and tertiary care) and is part of the sentinel sites for influenza surveillance in Japan.

### Sample collection and whole-genome sequencing

The residual nasopharyngeal swabs (NPS) from the RIDTs were temporarily stored at −20°C in the laboratory department of the hospital after being used for the RIDTs. The samples were transported to the Institute of Tropical Medicine at Nagasaki University within a week. The samples were stored in a deep freezer (−80°C) until further processed. Additional sociodemographic and clinical information on the patients was collected from the hospital database.

### RNA extraction and influenza virus detection

Viral nucleic acid was extracted directly from the NPS samples using a QIAamp viral RNA mini kit (QIAGEN Inc., Valencia, CA) following the manufacturer’s instructions. RNA was eluted to a final volume of 60 μL, aliquoted, and stored at −20°C for immediate use in reverse transcriptase-polymerase chain reaction (RT-PCR) or −80°C for long term storage. Multiplex RT- PCR assays were applied to screen influenza viruses (A and B) and other eleven other respiratory viruses. One Step RT-PCR Kit (QIAGEN Inc., Valencia, CA, USA) was used for RNA viruses, and GoTaq Flexi DNA Polymerase (Promega, San Luis Obispo, CA, USA) and PCR Nucleotide Mix (Promega, San Luis Obispo, CA, USA) were used for DNA viruses. The details of the multiplex PCR assay protocols were described elsewhere.^18^

### Multi-segment RT-PCR, library preparation and next-generation sequencing

For all RT-PCR-confirmed influenza A positive samples, all 8 segments of RT-PCR-confirmed influenza samples were amplified following the protocols of Zhou, B., et al. (2009)^19^ for influenza A. Each PCR product was purified again using Ampure XP beads (Beckman Coulter) according to the manufacturer’s instructions. The purity was then assessed with Agilent Technology 2100 Bioanalyzer using a High Sensitivity DNA chip and Qubit dsDNA HS Assay Kit (Life Technology). 1 ng of the DNA was used for library preparation (Nextera XT Kit, Illumina) following the manufacturer’s instructions (Illumina, CA, USA). The prepared library was sequenced on the MiSeq platform (Illumina) using a V2 2 × 250 bp reagents kit at Nagasaki University.

### Next-generation sequencing (NGS) data processing and genome assembly

A total of 254 samples were available for whole genome sequencing (supplementary figure S2). The sequencing output was uploaded to the INSaFLU,^20^ a web-based bioinformatic platform for influenza sequencing analysis. By providing raw sequence data, INSaFLU performs (i) quality check (using FastQC),^21^ and (ii) removal of adapter sequences and low-quality reads (using Trimmomatic)^22^. Trimmed and filtered sequences of less than 100 bp were discarded in downstream analysis. The quality-filtered sequences read were mapped to reference sequence A/H3N2_A_Perth_16_2009. Consensus for sequences with at least 10-fold coverage was then generated. The INSAFLU also performed the genome annotation (using Prokka)^23^.

### Phylogenetic analysis

To contextualize the Kamigoto samples within the landscape of viruses circulating in Japan and globally during the 2011/12 and 2012/2013 influenza seasons, we downloaded A/H3N2 influenza sequences from GISAID (https://www.gisaid.org) sampled between 1^st^ January 2011, and 31^st^ December 2013 for each individual segment (PB2, PB1, PA, HA, NP, NA, MP and NS) (as of 1^st^ January 2023). For each individual segment, sequence alignment was performed with MAFFT version 7 ^23^, and the alignments were visualized and manually edited in AliView. ^24^

Phylogenetic trees of each segment data were estimated using the maximum likelihood (ML) procedure in the IQTREE v2.0.7, ^25^ under the best-fitting model of nucleotide substitution TVM+F+I+I+R5, as determined by the ModelFinder option implemented in software. Branch supports were estimated by standard non-parametric bootstrap analysis with 100 replicates. TempEst v1.5.3^26^ was used to assess the presence of a molecular clock signal in the analyzed data, and linear regression of root-to-tip genetic distances against sampling dates was reconstructed. The outliers’ sequences and removed and reanalyzed.

We performed time-scaled phylogenies and root-to-tip linear regressions using TreeTime^27^ based on the maximum likelihood (ML) trees generated by IQTREE. The global sequences included in this analysis were Africa (n=39), Asia (n=180), Europe (n= 57), North America (n=365), Oceania (n=111), and South America (n=82). Time trees were derived utilizing the TreeTime function, incorporating up to 500 iterations, while considering covariation for estimating nucleotide substitution rates across the phylogeny. Automatic rerooting of the tree was performed to enhance clock-like signal. The ’mugration’ model within TreeTime was employed for ancestral state reconstruction, aiming to ascertain the probable migration patterns of the sampled viruses along the phylogeny, with genome sampling locations serving as discrete traits.

Additionally, time-resolved phylogenetic trees of HA segments were reconstructed through Bayesian inference using BEAST (v.10.1.4) ^28^ employing the Markov chain Monte Carlo (MCMC) method. A strict molecular clock with a Hasegawa-Kishino-Yano (HKY) nucleotide substitution model and a gamma-distributed model of among-site rate variation using four rate categories was used. A strict molecular clock was chosen based on the evolutionary distance in each data set, as estimated by TempEst^26^. The MCMC chain was run for at least 500 million generations, allowing 10% burn-in and sub-sampling every 50,000 steps. Convergence was confirmed when all posterior values provided effective sample sizes (ESS) greater than 200, with a 10% burn-in. The Tracer v1.5^29^ was used to detect the convergence. The maximum clade credibility (MCC) trees were summarized by TreeAnnotator v1.8.4 in the BEAST package.^30^ All phylogenetic trees were visualized and annotated in FigTree software^31^ and Inkscape^32^.

We also downloaded all publicly available full-length sequences of Japan for all eight influenza segments from the GISAID^32^ (n=88, as of April 2020). All eight influenza A(H3N2) coding sequences in the downloaded sequences from Japan and Kamigoto were concatenated into an alignment of 13,136 nucleotides in the order of the segment number (PB2-PB1-PA-HA-NP-NA-M-NS). The concatenated sequences were aligned using MAFFT version 7^23^ and the alignments were visualized and manually edited in AliView. ^24^ Maximum likelihood phylogenetic trees were reconstructed for the whole genome sequences of Japan and Kamigoto strains, as mentioned above.

## Data availability / Accession number(s)

All consensus sequences generated here have been submitted to the GISAID database. The details of isolate IDs and assigned accession numbers are listed in the supplementary data (Supplementary Table-1).

## Results

### Whole genome sequencing of A/H3N2 samples

Supplementary Figure 1 displays the epidemiological curve aggregated by reported RIDT positive cases and sequenced cases. A total of 254 samples were available for whole genome sequencing. Of these, 229 samples were characterized as A(H3N2) (29 samples from 2011/12 and 200 from 2012/2013 influenza season. Out of 229 A(H3N2) samples, 178 samples were successfully generated for WGS (see Supplementary Figure-S2 for a description of the sample selection process). Sequencing efficiency varied slightly, inversely with segment size, with smaller fragments such as the M segment generally displaying greater depth. The overview of the depth and length sequence coverage for each segment for all the samples is provided in supplementary Figure-S3.

The maximum likelihood phylogeny of WGS of A/H3N2 from Kamigoto island (Supplementary Figure S4) showed that Kamigoto sequences grouped closely together based on the year they were collected. The maximum likelihood phylogenetic analysis of the HA segment for Kamigoto strains revealed the prevalent circulation of clade 3C.2 in the island, and a few Kamigoto sequences (20) from 2012/13 were classified under 3C.3a (Figure 1). We also compared the other individual segments of Kamigoto strains (PB2, PB1, PA, NP, NA, MP and NS) to the publicly available strains from Japan and global sequences from GISAID as of the 1^st^ of December 2022 submission date (Supplementary Figure S-5 to S-12). We separately performed the phylogenetic analysis for each segment to avoid the reassortment bias between the segments.

**Figure 1:**
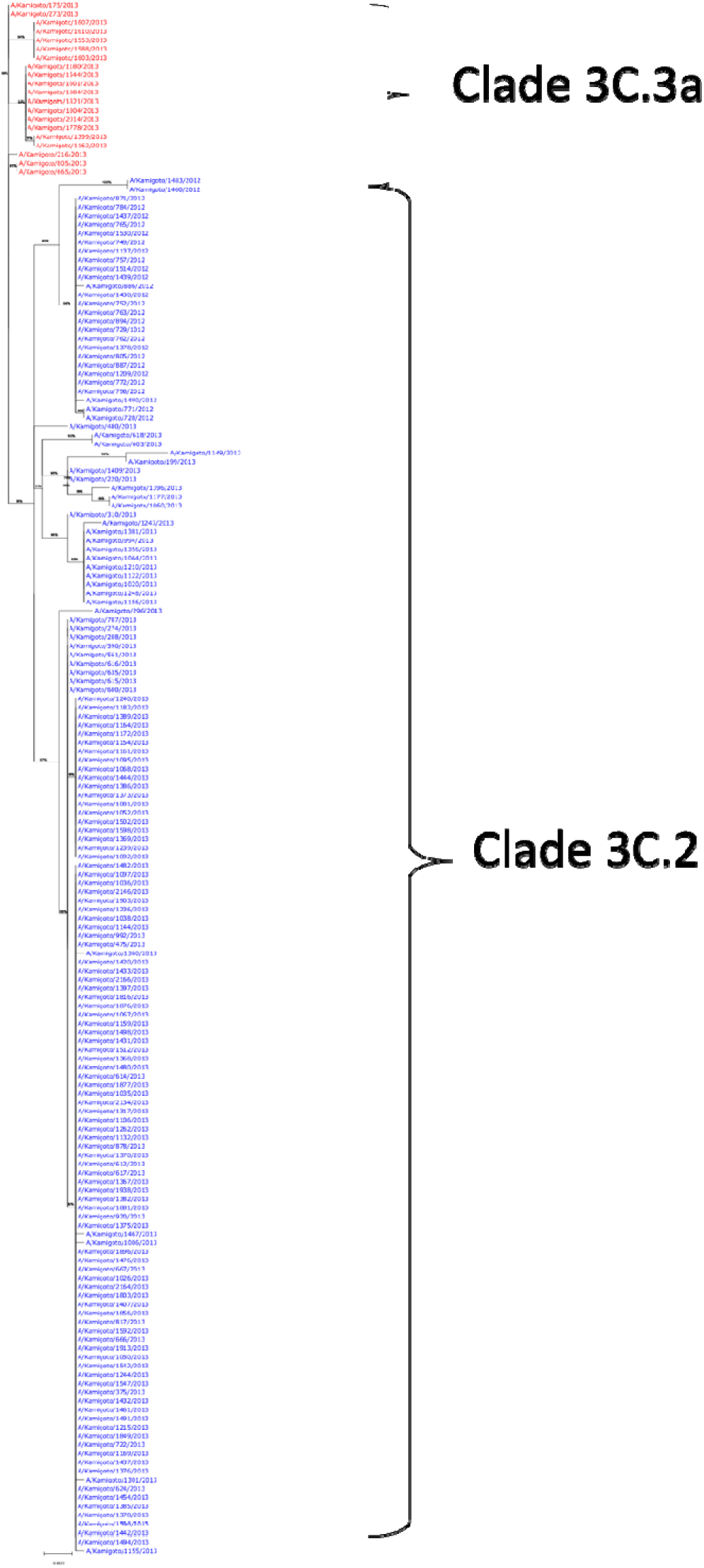
Maximum likelihood Phylogenetic tree of HA segments of A(H3N2) viruses from Kamigoto island, Japan

### Genetic diversity of A(H3N2) viruses isolated in Kamigoto island, Japan

We assessed how the HA segments of the sampled cases in Kamigoto compared to publicly available sequences from GISAID sampled globally between January 2011 and December 2013 by inferring a phylogenetic tree using the hemagglutinin (HA) sequences (Figure 2, supplementary figure S-13). The phylogenetic tree analysis showed that Kamigoto sequences grouped closely together based on the year they were collected. Within these, smaller and closely related sub-groups were identified. Groups of sequences are identified if (i) at least five sequences were grouped together, (ii) and with posterior branch support above 0.9, and (iii) from the same geographical location according to mugration modeling. Most of the Kamigoto sequences were classified into five distinct groups (Figure 2). The biggest group (Group 5) consists of 109 local sequences and 21 non-local sequences from mainland Japan (n=13), North America (n=6), Europe (n=1), and Asia (n=1). Sequences in Group 5 are further classified as 5A (28 sequences) and 5B (81 sequences), as these sequences are closer than other groups, but their date of coalescence is too far from their onset date of the cases, suggesting different introductions. The second biggest group (Group 1) was from the 2011/12 season, which consisted of 27 local sequences and 33 non-local sequences from mainland Japan (n=10), North America (n=12), Europe (n=3), Oceania (n=3), and Asia (n=5). The clustering of a large number of sequences (Group 5B, 5A and Group 1) on the phylogeny suggested that the majority of the cases sequenced in this study likely to be part of the local transmission chains. A few Kamigoto sequences (n=7) were isolated, without clustering with any other Kamigoto sequences. This suggested the presence of independent introductions that either did not cause onward transmission within the island or, alternatively, were not sampled in this study.

**Figure 2:**
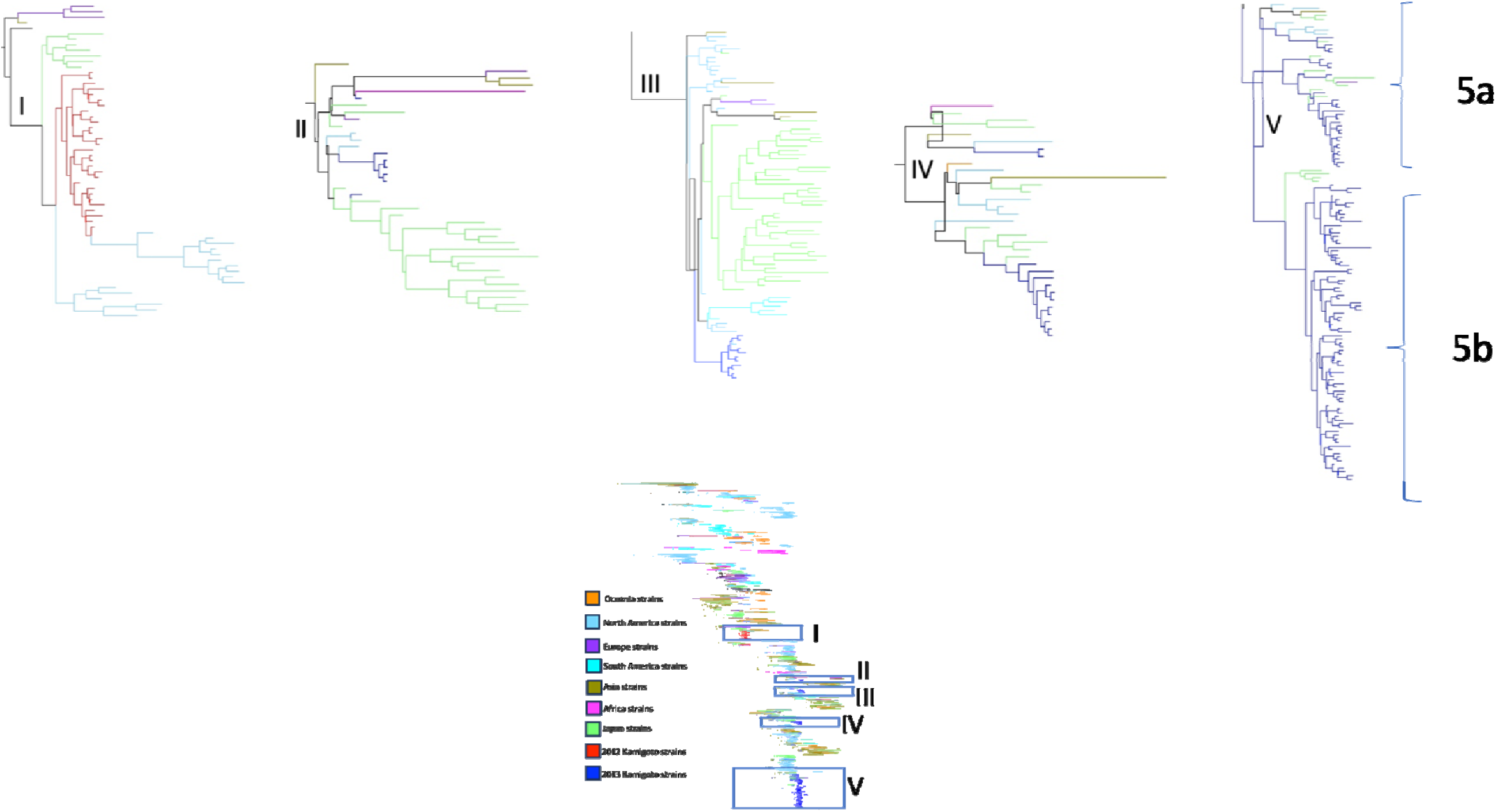
Clusters identified from time-resolved Phylogenetic tree of HA segments of A(H3N2) viruses from Kamigoto island, Japan and global sequences (GISAID) as of 1 December 2022 submission date.

### Local A(H3N2) clusters within the islands

2012/13 Kamigoto sequences were classified in five groups, which co-circulated during the same period, mainly between February and April 2013. (Figure 3). This suggested that at least five different importation events initiated the transmission and further drove local transmission. We then explored the geographical distribution of the Kamigoto strains within these groups (Figure 4). While location of the cases from Group 2 and Group 3 locations differed, Group 3, Group 4 and Group 5a had overlapping locations of the sequenced cases. The biggest local Kamigoto sequence group 5B was found to be distributed across the island (Figure 4).

**Figure 3:**
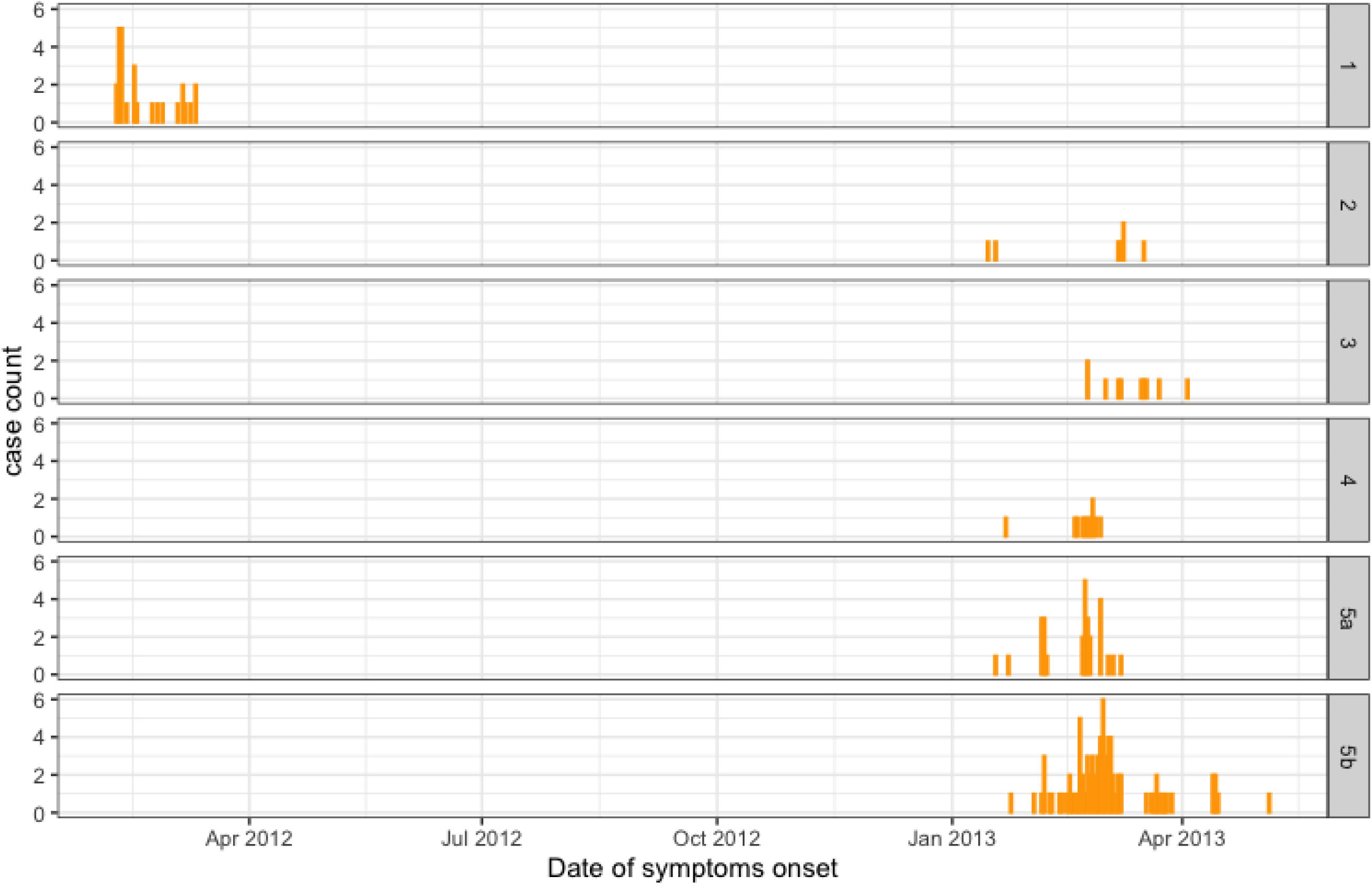
Distribution of sequenced H3N2 cases during the study period by group

**Figure 4:**
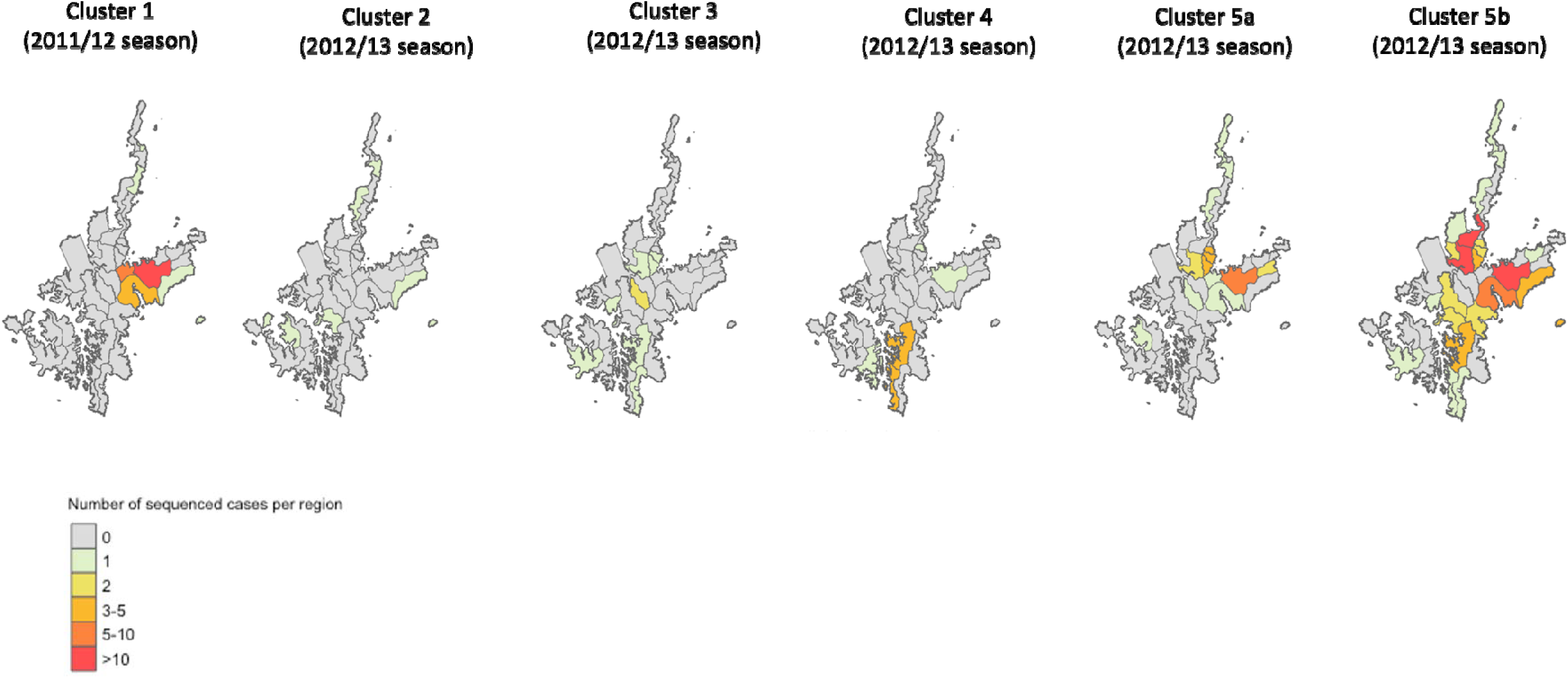
Geographic distribution of the clusters identified during 2011/12 and 2012/13 influenza seasons.

We further analyzed the biggest transmission group of Kamigoto sequences (Group 5A and 5B) to explore the spatio-temporal distribution patterns (Figure 5, Supplementary Figure S- 14). The spatio-temporal analysis of largest local sequences was predominantly characterized by the virus spreading from populous districts to less populous ones and between busier or more populous districts, such as Aogata-go (an urban area of the island) and Tainoura-go and Arikawa-go (port areas connecting to mainland Japan). The earliest sequenced case of Group 5A was reported on 18^th^ Januarys 2013. The number of cases increased in subsequent weeks in the same districts and further spread out to other districts in February and March. The earliest sequenced case of Group 5B was reported in January and the highest number of cases from this cluster was reported in February 2013. The outbreaks slowly declined from March onwards in both groups. RT-PCR confirmed Influenza positive surveillance data (supplementary Figure S-1) also showed February to April as peak of the season. A similar temporospatial distribution was also observed in local sequences cluster in group 1 (Supplementary Figure S-15).

**Figure 5:**
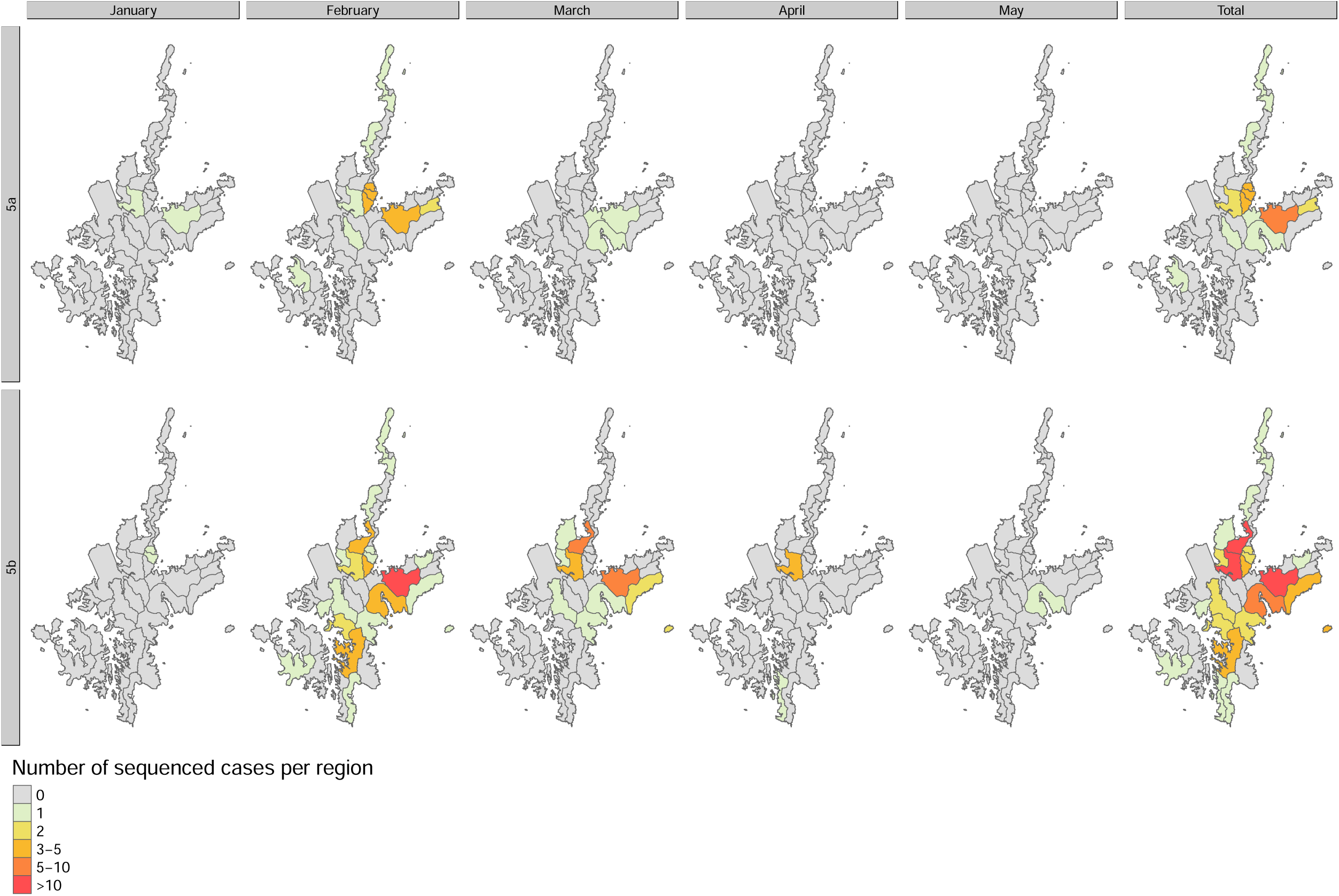
(A) Spatio-temporal distribution of the cluster 5A and 5B

## Discussion

In this study, we conducted a retrospective genomic characterization of seasonal influenza A/H3N2 viruses collected from Kamigoto island, Japan, during 2011/12 and 2012/13 influenza seasons. We extracted RNA from nasopharyngeal swab (NPS) samples that had been retrospectively collected during the 2011/12 and 2012/13 influenza seasons. We successfully sequenced the whole genomes of 178 A/H3N2 samples: 29 from the 2011/12 season and 149 from the 2012/13 season. This contribution is particularly valuable, as there are limited whole genome sequences (WGS) available from Japan for that time period (less than 10 in 2011/12 and zero in 2012/13 influenza season).

Regarding the time-resolved phylogeny, we identified more than six independent importations of A(H3N2) viruses into the Kamigoto island community during the study period and the continued local transmission and evolution of these viruses. Kamigoto is a semi-isolated island with only two ports linking it to mainland Japan. The phylogeny identified that the Kamigoto sequences were closely related to strains from mainland Japan, and local sequences in each group were observed to be closely clustered with strains from mainland Japan. This contrasts with findings from studies conducted in a Kenyan city^24^ and Basel city in Switzerland^25^. In those studies, strains from Kenya and Basel displayed close genetic relations to global strains, reflecting the high connectivity of these urban centers with other parts of the world. Such differences highlight the significance of understanding influenza dynamics at the local level since transmission patterns can vary based on geographical factors. Interestingly, our study revealed a few potential international importation events where the Kamigoto strains closely resembled those from outside Japan in the phylogenetic analysis. This underscores the importance of regularly testing visitors and returning residents, especially during influenza. Low density of sequence numbers available from the southern part of Japan during the same period (only GISAID) may also be another reason explaining the close relatedness of the Kamigoto strains to outside Japan.

We found Kamigoto strains clustered in more than six different groups in the phylogenetic tree. Notably, Kamigoto sequence clusters in 2012/13 were co-circulating during the same period, especially between February and April 2013, indicating that different importation events fueled the transmission of influenza in each group. In absence of genomic surveillance, cases circulating simultaneously may be inaccurately classified into the same cluster using only epidemiological data, which would lead to underestimations of the importation risks, and potential overestimations of the risk of transmission. Integrating sequence data with epidemiological information, provided the geographical distribution of cases in each group. We identified five sequences that did not closely relate to any other sequences in our study that might represent independent introductions. It’s possible that subsequent transmission for these outliers either didn’t occur or cases arising from them weren’t available for sequencing to be included in this study. In this study, we were able to sequence only 254 samples out of 538 RT-PCR confirmed influenza cases reported (Suppplementary Figure S2).

The largest cluster of local sequences (Group 5B) contained 81 sequences, all from Kamigoto, suggesting that they all belonged to the same local transmission chain. The second largest group (Group 5A) included 28 Kamigoto sequences and was found to be clustered with sequences from mainland Japan (n=9) and other parts of the world. Kamigoto sequences in Group 5A and 5B identified on the phylogeny suggested repeated importations of similar strains, potentially circulating simultaneously outside Kamigoto, primarily from mainland Japan. This could be due to concurrent transmission in mainland Japan and Kamigoto, with repeated introductions of the same strain. The finding of the spatial distribution of the sequences in Group 5B align with our earlier epidemiological study,^2^ which identified districts with connections outside the island and high population density as the main drivers of influenza transmission patterns at the local community level.

Although the study used samples that were collected more than 9 years ago, a good viral yield was retrieved by direct RNA extraction from the samples. This is a benefit of sequencing directly from the samples (rather than passaging or isolation) and storage with optimal temperature for long duration. Only few sequences were available from the Kyuushuu regions (where Kamigoto is located) prior to this project. The sequences collected, shared, and analysed in this paper therefore greatly improve our understanding and knowledge of influenza phylogeny in Japan.

The sequenced samples were dependent on the sample viral load and cDNA concentration to pass the sequencing quality and thus it limits the representativeness of the data in reconstruction of the influenza transmission. However, the sequenced samples were roughly proportional to the reported RIDT-confirmed influenza cases. The number of sequences generated in this study were relatively low compared to the reported number of influenza cases in that period (1,009 and 1,053 RIDT cases were reported in 2011/12 and 2012/13, Supplementary Figure 1). An increased sequencing frequency may capture more introduction events and clusters. Finally, the study was part of the surveillance system, and thus data were collected during the influenzas season (October-June) rather than the whole year. This limits our study’s assessment of year-round influenza transmission patterns within the islands. Conclusion

This analysis helps to better understand the transmission patterns of seasonal influenza because it was conducted in rural setting such as Kamigoto island with a semi-isolated population in Japan. Influenza A(H3N2) virus epidemics in Kamigoto island were marked by multiple introductions and fueled by local transmission. A closer examination of local transmission through genomic data indicates concurrent independent introduction events and local proliferation. These might be misinterpreted as part of the same cluster without sequencing data. However, the results of this study are based on a two-years analysis of influenza sequences from the island, thus repeated analyzes for different influenza seasons and geographic locations will help us better understand detailed transmission patterns.

## Ethics

The research was approved by the institutional review boards of Kamigoto Hospital, Nagasaki University Research Ethics Committee (reference number 200619236), and the London School of Hygiene and Tropical Medicine Research Ethics Committee (reference number 26706). Both Nagasaki University and London School of Hygiene and Tropical Medicine granted waivers for obtaining informed consent due to the nature of this retrospective study and the preserved anonymity of patients.

## Data Availability

All data produced are available online at GISAID

https://gisaid.org

## Supplementary data

Supplementary data are available at online.

## Acknowledgements

We would like to thank those who participated in the study and the staff at Kamigoto Hospital for their work in collecting and managing data.

