## Supplementary for "Phylogenetic study of local patterns influenza A(H3N2) virus transmission in a semi-isolated population in a remote island in Japan between 2011-2013"

Table S-1: GISAID accession number of sequences produced in this study

| **Isolate_Name** | **Isolate_Id** | **PB2** | **PB1** | **PA** | **HA** | **NP** | **NA** | **MP** | **NS** |
| --- | --- | --- | --- | --- | --- | --- | --- | --- | --- |
| A/Kamigoto/728/2012 | EPI_ISL_17103063 | EPI2449327 | EPI2449328 | EPI2449329 | EPI2449330 | EPI2449331 | EPI2449332 | EPI2449333 | EPI2449334 |
| A/Kamigoto/199/2013 | EPI_ISL_17092317 | EPI2440111 | EPI2440227 | EPI2440228 | EPI2440229 | EPI2440230 | EPI2440231 | EPI2440232 | EPI2440233 |
| A/Kamigoto/1447/2013 | EPI_ISL_18002728 | EPI2635351 | EPI2635352 | EPI2635353 | EPI2635354 | EPI2635355 | EPI2635356 | EPI2635357 | EPI2635358 |
| A/Kamigoto/1454/2013 | EPI_ISL_18002450 | EPI2635343 | EPI2635344 | EPI2635345 | EPI2635346 | EPI2635347 | EPI2635348 | EPI2635349 | EPI2635350 |
| A/Kamigoto/1461/2013 | EPI_ISL_18002449 | EPI2635335 | EPI2635336 | EPI2635337 | EPI2635338 | EPI2635339 | EPI2635340 | EPI2635341 | EPI2635342 |
| A/Kamigoto/1444/2013 | EPI_ISL_18002434 | EPI2635327 | EPI2635328 | EPI2635329 | EPI2635330 | EPI2635331 | EPI2635332 | EPI2635333 | EPI2635334 |
| A/Kamigoto/1467/2013 | EPI_ISL_18002433 | EPI2635319 | EPI2635320 | EPI2635321 | EPI2635322 | EPI2635323 | EPI2635324 | EPI2635325 | EPI2635326 |
| A/Kamigoto/1442/2013 | EPI_ISL_18002432 | EPI2635311 | EPI2635312 | EPI2635313 | EPI2635314 | EPI2635315 | EPI2635316 | EPI2635317 | EPI2635318 |
| A/Kamigoto/1476/2013 | EPI_ISL_18001909 | EPI2635303 | EPI2635304 | EPI2635305 | EPI2635306 | EPI2635307 | EPI2635308 | EPI2635309 | EPI2635310 |
| A/Kamigoto/1480/2013 | EPI_ISL_18001908 | EPI2635295 | EPI2635296 | EPI2635297 | EPI2635298 | EPI2635299 | EPI2635300 | EPI2635301 | EPI2635302 |
| A/Kamigoto/1437/2013 | EPI_ISL_18001895 | EPI2635239 | EPI2635240 | EPI2635241 | EPI2635242 | EPI2635253 | EPI2635265 | EPI2635278 | EPI2635283 |
| A/Kamigoto/1433/2013 | EPI_ISL_18001894 | EPI2635231 | EPI2635232 | EPI2635233 | EPI2635234 | EPI2635235 | EPI2635236 | EPI2635237 | EPI2635238 |
| A/Kamigoto/1432/2013 | EPI_ISL_18001808 | EPI2635135 | EPI2635136 | EPI2635146 | EPI2635161 | EPI2635174 | EPI2635186 | EPI2635197 | EPI2635211 |
| A/Kamigoto/1431/2013 | EPI_ISL_18001804 | EPI2635127 | EPI2635128 | EPI2635129 | EPI2635130 | EPI2635131 | EPI2635132 | EPI2635133 | EPI2635134 |
| A/Kamigoto/1420/2013 | EPI_ISL_18001803 | EPI2635119 | EPI2635120 | EPI2635121 | EPI2635122 | EPI2635123 | EPI2635124 | EPI2635125 | EPI2635126 |
| A/Kamigoto/1482/2013 | EPI_ISL_18001802 | EPI2635111 | EPI2635112 | EPI2635113 | EPI2635114 | EPI2635115 | EPI2635116 | EPI2635117 | EPI2635118 |
| A/Kamigoto/1491/2013 | EPI_ISL_18001801 | EPI2635076 | EPI2635094 | EPI2635105 | EPI2635106 | EPI2635107 | EPI2635108 | EPI2635109 | EPI2635110 |
| A/Kamigoto/1494/2013 | EPI_ISL_18001747 | EPI2634970 | EPI2634971 | EPI2634972 | EPI2634973 | EPI2634974 | EPI2634975 | EPI2634976 | EPI2634977 |
| A/Kamigoto/1498/2013 | EPI_ISL_18001746 | EPI2634962 | EPI2634963 | EPI2634964 | EPI2634965 | EPI2634966 | EPI2634967 | EPI2634968 | EPI2634969 |
| A/Kamigoto/1502/2013 | EPI_ISL_18001715 | EPI2634954 | EPI2634955 | EPI2634956 | EPI2634957 | EPI2634958 | EPI2634959 | EPI2634960 | EPI2634961 |
| A/Kamigoto/1512/2013 | EPI_ISL_18001714 | EPI2634946 | EPI2634947 | EPI2634948 | EPI2634949 | EPI2634950 | EPI2634951 | EPI2634952 | EPI2634953 |
| A/Kamigoto/1542/2013 | EPI_ISL_18001713 | EPI2634937 | EPI2634938 | EPI2634939 | EPI2634940 | EPI2634941 | EPI2634942 | EPI2634943 | EPI2634944 |
| A/Kamigoto/1544/2013 | EPI_ISL_18001707 | EPI2634922 | EPI2634923 | EPI2634924 | EPI2634925 | EPI2634926 | EPI2634927 | EPI2634928 | EPI2634929 |
| A/Kamigoto/1547/2013 | EPI_ISL_18001706 | EPI2634865 | EPI2634887 | EPI2634908 | EPI2634917 | EPI2634918 | EPI2634919 | EPI2634920 | EPI2634921 |
| A/Kamigoto/1553/2013 | EPI_ISL_18001705 | EPI2634844 | EPI2634845 | EPI2634846 | EPI2634847 | EPI2634848 | EPI2634849 | EPI2634850 | EPI2634851 |
| A/Kamigoto/1588/2013 | EPI_ISL_18001704 | EPI2634835 | EPI2634837 | EPI2634838 | EPI2634839 | EPI2634840 | EPI2634841 | EPI2634842 | EPI2634843 |
| A/Kamigoto/1592/2013 | EPI_ISL_18001703 | EPI2634778 | EPI2634798 | EPI2634819 | EPI2634830 | EPI2634831 | EPI2634832 | EPI2634833 | EPI2634834 |
| A/Kamigoto/1596/2013 | EPI_ISL_18001702 | EPI2634504 | EPI2634523 | EPI2634543 | EPI2634563 | EPI2634578 | EPI2634600 | EPI2634618 | EPI2634641 |
| A/Kamigoto/1598/2013 | EPI_ISL_18001701 | EPI2634425 | EPI2634426 | EPI2634427 | EPI2634428 | EPI2634429 | EPI2634430 | EPI2634431 | EPI2634432 |
| A/Kamigoto/1601/2013 | EPI_ISL_18001686 | EPI2634336 | EPI2634350 | EPI2634362 | EPI2634374 | EPI2634387 | EPI2634401 | EPI2634412 | EPI2634415 |
| A/Kamigoto/1603/2013 | EPI_ISL_18001652 | EPI2634085 | EPI2634099 | EPI2634110 | EPI2634124 | EPI2634134 | EPI2634146 | EPI2634156 | EPI2634174 |
| A/Kamigoto/1607/2013 | EPI_ISL_18001626 | EPI2633893 | EPI2633905 | EPI2633917 | EPI2633935 | EPI2633946 | EPI2633958 | EPI2633971 | EPI2633981 |
| A/Kamigoto/1610/2013 | EPI_ISL_18001610 | EPI2633719 | EPI2633745 | EPI2633763 | EPI2633764 | EPI2633765 | EPI2633766 | EPI2633767 | EPI2633775 |
| A/Kamigoto/1778/2013 | EPI_ISL_18001609 | EPI2633703 | EPI2633704 | EPI2633705 | EPI2633706 | EPI2633707 | EPI2633708 | EPI2633709 | EPI2633710 |
| A/Kamigoto/1803/2013 | EPI_ISL_18001608 | EPI2633695 | EPI2633696 | EPI2633697 | EPI2633698 | EPI2633699 | EPI2633700 | EPI2633701 | EPI2633702 |
| A/Kamigoto/1804/2013 | EPI_ISL_18001607 | EPI2633687 | EPI2633688 | EPI2633689 | EPI2633690 | EPI2633691 | EPI2633692 | EPI2633693 | EPI2633694 |
| A/Kamigoto/1816/2013 | EPI_ISL_18001606 | EPI2633679 | EPI2633680 | EPI2633681 | EPI2633682 | EPI2633683 | EPI2633684 | EPI2633685 | EPI2633686 |
| A/Kamigoto/1821/2013 | EPI_ISL_18001522 | EPI2633671 | EPI2633672 | EPI2633673 | EPI2633674 | EPI2633675 | EPI2633676 | EPI2633677 | EPI2633678 |
| A/Kamigoto/1849/2013 | EPI_ISL_18001521 | EPI2633663 | EPI2633664 | EPI2633665 | EPI2633666 | EPI2633667 | EPI2633668 | EPI2633669 | EPI2633670 |
| A/Kamigoto/1856/2013 | EPI_ISL_18001520 | EPI2633655 | EPI2633656 | EPI2633657 | EPI2633658 | EPI2633659 | EPI2633660 | EPI2633661 | EPI2633662 |
| A/Kamigoto/1876/2013 | EPI_ISL_18001519 | NA | NA | EPI2633649 | EPI2633650 | EPI2633651 | EPI2633652 | EPI2633653 | EPI2633654 |
| A/Kamigoto/1877/2013 | EPI_ISL_18001518 | EPI2633641 | EPI2633642 | EPI2633643 | EPI2633644 | EPI2633645 | EPI2633646 | EPI2633647 | EPI2633648 |
| A/Kamigoto/1881/2013 | EPI_ISL_18001517 | EPI2633633 | EPI2633634 | EPI2633635 | EPI2633636 | EPI2633637 | EPI2633638 | EPI2633639 | EPI2633640 |
| A/Kamigoto/1884/2013 | EPI_ISL_18001516 | EPI2633625 | EPI2633626 | EPI2633627 | EPI2633628 | EPI2633629 | EPI2633630 | EPI2633631 | EPI2633632 |
| A/Kamigoto/1896/2013 | EPI_ISL_18001514 | EPI2633617 | EPI2633618 | EPI2633619 | EPI2633620 | EPI2633621 | EPI2633622 | EPI2633623 | EPI2633624 |
| A/Kamigoto/1903/2013 | EPI_ISL_18001513 | EPI2633301 | EPI2633302 | EPI2633303 | EPI2633304 | EPI2633305 | EPI2633306 | EPI2633307 | EPI2633311 |
| A/Kamigoto/1913/2013 | EPI_ISL_18001512 | EPI2633293 | EPI2633294 | EPI2633295 | EPI2633296 | EPI2633297 | EPI2633298 | EPI2633299 | EPI2633300 |
| A/Kamigoto/1938/2013 | EPI_ISL_18001511 | EPI2633285 | EPI2633286 | EPI2633287 | EPI2633288 | EPI2633289 | EPI2633290 | EPI2633291 | EPI2633292 |
| A/Kamigoto/2014/2013 | EPI_ISL_18001510 | EPI2633277 | EPI2633278 | EPI2633279 | EPI2633280 | EPI2633281 | EPI2633282 | EPI2633283 | EPI2633284 |
| A/Kamigoto/2134/2013 | EPI_ISL_18001486 | NA | NA | NA | EPI2633272 | EPI2633273 | EPI2633274 | EPI2633275 | EPI2633276 |
| A/Kamigoto/2146/2013 | EPI_ISL_18000339 | NA | NA | NA | EPI2632321 | EPI2632322 | EPI2632323 | EPI2632324 | EPI2632325 |
| A/Kamigoto/2164/2013 | EPI_ISL_18000338 | NA | EPI2632314 | EPI2632315 | EPI2632316 | EPI2632317 | EPI2632318 | EPI2632319 | EPI2632320 |
| A/Kamigoto/2166/2013 | EPI_ISL_18000336 | NA | NA | NA | EPI2632301 | EPI2632302 | EPI2632304 | EPI2632306 | EPI2632308 |
| A/Kamigoto/1419/2013 | EPI_ISL_17786021 | NA | NA | NA | EPI2589364 | EPI2589365 | EPI2589366 | EPI2589367 | EPI2589368 |
| A/Kamigoto/1409/2013 | EPI_ISL_17785996 | EPI2589356 | EPI2589357 | EPI2589358 | EPI2589359 | EPI2589360 | EPI2589361 | EPI2589362 | EPI2589363 |
| A/Kamigoto/1407/2013 | EPI_ISL_17785995 | EPI2589348 | EPI2589349 | EPI2589350 | EPI2589351 | EPI2589352 | EPI2589353 | EPI2589354 | EPI2589355 |
| A/Kamigoto/1399/2013 | EPI_ISL_17785743 | EPI2589340 | EPI2589341 | EPI2589342 | EPI2589343 | EPI2589344 | EPI2589345 | EPI2589346 | EPI2589347 |
| A/Kamigoto/1397/2013 | EPI_ISL_17785739 | EPI2589324 | EPI2589325 | EPI2589326 | EPI2589327 | EPI2589328 | EPI2589329 | EPI2589330 | EPI2589331 |
| A/Kamigoto/1396/2013 | EPI_ISL_17785738 | EPI2589316 | EPI2589317 | EPI2589318 | EPI2589319 | EPI2589320 | EPI2589321 | EPI2589322 | EPI2589323 |
| A/Kamigoto/1389/2013 | EPI_ISL_17785724 | EPI2589308 | EPI2589309 | EPI2589310 | EPI2589311 | EPI2589312 | EPI2589313 | EPI2589314 | EPI2589315 |
| A/Kamigoto/1386/2013 | EPI_ISL_17785723 | EPI2589228 | EPI2589229 | EPI2589230 | EPI2589231 | EPI2589232 | EPI2589233 | EPI2589234 | EPI2589235 |
| A/Kamigoto/1385/2013 | EPI_ISL_17785722 | EPI2589220 | EPI2589221 | EPI2589222 | EPI2589223 | EPI2589224 | EPI2589225 | EPI2589226 | EPI2589227 |
| A/Kamigoto/1382/2013 | EPI_ISL_17785719 | EPI2589212 | EPI2589213 | EPI2589214 | EPI2589215 | EPI2589216 | EPI2589217 | EPI2589218 | EPI2589219 |
| A/Kamigoto/1381/2013 | EPI_ISL_17785716 | EPI2589204 | EPI2589205 | EPI2589206 | EPI2589207 | EPI2589208 | EPI2589209 | EPI2589210 | EPI2589211 |
| A/Kamigoto/1378/2013 | EPI_ISL_17785715 | EPI2589196 | EPI2589197 | EPI2589198 | EPI2589199 | EPI2589200 | EPI2589201 | EPI2589202 | EPI2589203 |
| A/Kamigoto/1376/2013 | EPI_ISL_17785627 | EPI2588519 | EPI2588520 | EPI2588521 | EPI2588522 | EPI2588523 | EPI2588524 | NA | EPI2588525 |
| A/Kamigoto/1375/2013 | EPI_ISL_17785624 | EPI2588511 | EPI2588512 | EPI2588513 | EPI2588514 | EPI2588515 | EPI2588516 | EPI2588517 | EPI2588518 |
| A/Kamigoto/1373/2013 | EPI_ISL_17785619 | EPI2588503 | EPI2588504 | EPI2588505 | EPI2588506 | EPI2588507 | EPI2588508 | EPI2588509 | EPI2588510 |
| A/Kamigoto/1370/2013 | EPI_ISL_17785617 | EPI2588495 | EPI2588496 | EPI2588497 | EPI2588498 | EPI2588499 | EPI2588500 | EPI2588501 | EPI2588502 |
| A/Kamigoto/1369/2013 | EPI_ISL_17785616 | EPI2588487 | EPI2588488 | EPI2588489 | EPI2588490 | EPI2588491 | EPI2588492 | EPI2588493 | EPI2588494 |
| A/Kamigoto/1368/2013 | EPI_ISL_17785615 | EPI2588479 | EPI2588480 | EPI2588481 | EPI2588482 | EPI2588483 | EPI2588484 | EPI2588485 | EPI2588486 |
| A/Kamigoto/1367/2013 | EPI_ISL_17785614 | EPI2588471 | EPI2588472 | EPI2588473 | EPI2588474 | EPI2588475 | EPI2588476 | EPI2588477 | EPI2588478 |
| A/Kamigoto/1360/2013 | EPI_ISL_17785613 | NA | NA | EPI2588465 | EPI2588466 | EPI2588467 | EPI2588468 | EPI2588469 | EPI2588470 |
| A/Kamigoto/1356/2013 | EPI_ISL_17761517 | NA | NA | NA | EPI2582636 | EPI2582637 | EPI2582638 | EPI2582639 | EPI2582640 |
| A/Kamigoto/1317/2013 | EPI_ISL_17761516 | EPI2582626 | EPI2582627 | EPI2582628 | EPI2582629 | EPI2582630 | EPI2582631 | EPI2582632 | EPI2582633 |
| A/Kamigoto/1301/2013 | EPI_ISL_17739156 | EPI2580996 | EPI2580997 | EPI2580998 | EPI2580999 | EPI2581000 | EPI2581001 | EPI2581002 | EPI2581003 |
| A/Kamigoto/1262/2013 | EPI_ISL_17738971 | EPI2580988 | EPI2580989 | EPI2580990 | EPI2580991 | EPI2580992 | EPI2580993 | EPI2580994 | EPI2580995 |
| A/Kamigoto/1248/2013 | EPI_ISL_17738944 | EPI2580980 | EPI2580981 | EPI2580982 | EPI2580983 | EPI2580984 | EPI2580985 | EPI2580986 | EPI2580987 |
| A/Kamigoto/1247/2013 | EPI_ISL_17730095 | NA | NA | NA | EPI2579590 | EPI2579591 | EPI2579592 | EPI2579593 | EPI2579594 |
| A/Kamigoto/1244/2013 | EPI_ISL_17730092 | EPI2579582 | EPI2579583 | EPI2579584 | EPI2579585 | EPI2579586 | EPI2579587 | EPI2579588 | EPI2579589 |
| A/Kamigoto/1240/2013 | EPI_ISL_17730088 | EPI2579574 | EPI2579575 | EPI2579576 | EPI2579577 | EPI2579578 | EPI2579579 | EPI2579580 | EPI2579581 |
| A/Kamigoto/1239/2013 | EPI_ISL_17730086 | EPI2579566 | EPI2579567 | EPI2579568 | EPI2579569 | EPI2579570 | EPI2579571 | EPI2579572 | EPI2579573 |
| A/Kamigoto/1236/2013 | EPI_ISL_17730085 | EPI2579558 | EPI2579559 | EPI2579560 | EPI2579561 | EPI2579562 | EPI2579563 | EPI2579564 | EPI2579565 |
| A/Kamigoto/1215/2013 | EPI_ISL_17730082 | EPI2579542 | EPI2579543 | EPI2579544 | EPI2579545 | EPI2579546 | EPI2579547 | EPI2579548 | EPI2579549 |
| A/Kamigoto/1210/2013 | EPI_ISL_17730079 | EPI2579534 | EPI2579535 | EPI2579536 | EPI2579537 | EPI2579538 | EPI2579539 | EPI2579540 | EPI2579541 |
| A/Kamigoto/1182/2013 | EPI_ISL_17730075 | EPI2579526 | EPI2579527 | EPI2579528 | EPI2579529 | EPI2579530 | EPI2579531 | EPI2579532 | EPI2579533 |
| A/Kamigoto/1180/2013 | EPI_ISL_17730072 | EPI2579517 | EPI2579518 | EPI2579519 | EPI2579520 | EPI2579521 | EPI2579523 | EPI2579524 | EPI2579525 |
| A/Kamigoto/1177/2013 | EPI_ISL_17730069 | EPI2579508 | EPI2579509 | EPI2579511 | EPI2579512 | EPI2579513 | EPI2579514 | EPI2579515 | EPI2579516 |
| A/Kamigoto/1172/2013 | EPI_ISL_17730068 | EPI2579500 | EPI2579501 | EPI2579502 | EPI2579503 | EPI2579504 | EPI2579505 | EPI2579506 | EPI2579507 |
| A/Kamigoto/1169/2013 | EPI_ISL_17730067 | EPI2579492 | EPI2579493 | EPI2579494 | EPI2579495 | EPI2579496 | EPI2579497 | EPI2579498 | EPI2579499 |
| A/Kamigoto/1164/2013 | EPI_ISL_17730066 | EPI2579484 | EPI2579485 | EPI2579486 | EPI2579487 | EPI2579488 | EPI2579489 | EPI2579490 | EPI2579491 |
| A/Kamigoto/1162/2013 | EPI_ISL_17730065 | EPI2579476 | EPI2579477 | EPI2579478 | EPI2579479 | EPI2579480 | EPI2579481 | EPI2579482 | EPI2579483 |
| A/Kamigoto/1161/2013 | EPI_ISL_17730047 | EPI2579468 | EPI2579469 | EPI2579470 | EPI2579471 | EPI2579472 | EPI2579473 | EPI2579474 | EPI2579475 |
| A/Kamigoto/1159/2013 | EPI_ISL_17730041 | EPI2579461 | NA | EPI2579462 | EPI2579463 | EPI2579464 | EPI2579465 | EPI2579466 | EPI2579467 |
| A/Kamigoto/1156/2013 | EPI_ISL_17726035 | EPI2579164 | EPI2579165 | EPI2579166 | EPI2579167 | EPI2579168 | EPI2579169 | EPI2579170 | EPI2579171 |
| A/Kamigoto/1155/2013 | EPI_ISL_17692316 | EPI2563277 | EPI2563279 | EPI2563278 | EPI2579159 | EPI2579160 | EPI2579161 | EPI2579162 | EPI2579163 |
| A/Kamigoto/1154/2013 | EPI_ISL_17692307 | EPI2563266 | EPI2563269 | EPI2563268 | EPI2563270 | EPI2563271 | EPI2563272 | EPI2563273 | EPI2563274 |
| A/Kamigoto/1149/2013 | EPI_ISL_17692279 | EPI2563258 | EPI2563259 | EPI2563260 | EPI2563261 | EPI2563262 | EPI2563263 | EPI2563264 | EPI2563265 |
| A/Kamigoto/1144/2013 | EPI_ISL_17692278 | EPI2563250 | EPI2563251 | EPI2563252 | EPI2563253 | EPI2563254 | EPI2563255 | EPI2563256 | EPI2563257 |
| A/Kamigoto/1132/2013 | EPI_ISL_17692276 | EPI2563242 | EPI2563243 | EPI2563244 | EPI2563245 | EPI2563246 | EPI2563247 | EPI2563248 | EPI2563249 |
| A/Kamigoto/1122/2013 | EPI_ISL_17692275 | EPI2563234 | EPI2563235 | EPI2563236 | EPI2563237 | EPI2563238 | EPI2563239 | EPI2563240 | EPI2563241 |
| A/Kamigoto/1108/2013 | EPI_ISL_17692274 | NA | NA | NA | EPI2563230 | NA | EPI2563231 | EPI2563232 | EPI2563233 |
| A/Kamigoto/1106/2013 | EPI_ISL_17692272 | EPI2563222 | EPI2563223 | EPI2563224 | EPI2563225 | EPI2563226 | EPI2563227 | EPI2563228 | EPI2563229 |
| A/Kamigoto/1097/2013 | EPI_ISL_17692271 | EPI2579550 | EPI2579551 | EPI2579552 | EPI2579553 | EPI2579554 | EPI2579555 | EPI2579556 | EPI2579557 |
| A/Kamigoto/1095/2013 | EPI_ISL_17692267 | EPI2563214 | EPI2563215 | EPI2563216 | EPI2563217 | EPI2563218 | EPI2563219 | EPI2563220 | EPI2563221 |
| A/Kamigoto/1092/2013 | EPI_ISL_17692266 | EPI2563206 | EPI2563207 | EPI2563208 | EPI2563209 | EPI2563210 | EPI2563211 | EPI2563212 | EPI2563213 |
| A/Kamigoto/1081/2013 | EPI_ISL_17692265 | EPI2563198 | EPI2563199 | EPI2563200 | EPI2563201 | EPI2563202 | EPI2563203 | EPI2563204 | EPI2563205 |
| A/Kamigoto/1068/2013 | EPI_ISL_17692264 | EPI2563190 | EPI2563191 | EPI2563192 | EPI2563193 | EPI2563194 | EPI2563195 | EPI2563196 | EPI2563197 |
| A/Kamigoto/1067/2013 | EPI_ISL_17692257 | EPI2563182 | EPI2563183 | EPI2563184 | EPI2563185 | EPI2563186 | EPI2563187 | EPI2563188 | EPI2563189 |
| A/Kamigoto/1064/2013 | EPI_ISL_17692212 | EPI2563174 | EPI2563175 | EPI2563176 | EPI2563177 | EPI2563178 | EPI2563179 | EPI2563180 | EPI2563181 |
| A/Kamigoto/1060/2013 | EPI_ISL_17692112 | EPI2563156 | EPI2563157 | EPI2563158 | EPI2563159 | EPI2563160 | EPI2563161 | EPI2563162 | EPI2563163 |
| A/Kamigoto/1052/2013 | EPI_ISL_17692053 | EPI2563148 | EPI2563149 | EPI2563150 | EPI2563151 | EPI2563152 | EPI2563153 | EPI2563154 | EPI2563155 |
| A/Kamigoto/1050/2013 | EPI_ISL_17692052 | EPI2563140 | EPI2563141 | EPI2563142 | EPI2563143 | EPI2563144 | EPI2563145 | EPI2563146 | EPI2563147 |
| A/Kamigoto/1038/2013 | EPI_ISL_17692051 | EPI2563132 | EPI2563133 | EPI2563134 | EPI2563135 | EPI2563136 | EPI2563137 | EPI2563138 | EPI2563139 |
| A/Kamigoto/1036/2013 | EPI_ISL_17692004 | EPI2563032 | EPI2563033 | EPI2563034 | EPI2563035 | EPI2563036 | EPI2563037 | EPI2563038 | EPI2563039 |
| A/Kamigoto/1035/2013 | EPI_ISL_17691998 | EPI2563024 | EPI2563025 | EPI2563026 | EPI2563027 | EPI2563028 | EPI2563029 | EPI2563030 | EPI2563031 |
| A/Kamigoto/1026/2013 | EPI_ISL_17691980 | EPI2563016 | EPI2563017 | EPI2563018 | EPI2563019 | EPI2563020 | EPI2563021 | EPI2563022 | EPI2563023 |
| A/Kamigoto/1020/2013 | EPI_ISL_17691968 | EPI2563001 | EPI2563004 | EPI2563006 | EPI2563008 | EPI2563009 | EPI2563011 | EPI2563012 | EPI2563014 |
| A/Kamigoto/1006/2013 | EPI_ISL_17691966 | EPI2562992 | EPI2562993 | EPI2562994 | EPI2562995 | EPI2562996 | EPI2562997 | EPI2562998 | EPI2562999 |
| A/Kamigoto/994/2013 | EPI_ISL_17691965 | EPI2562984 | EPI2562985 | EPI2562986 | EPI2562987 | EPI2562988 | EPI2562989 | EPI2562990 | EPI2562991 |
| A/Kamigoto/992/2013 | EPI_ISL_17689263 | EPI2562656 | EPI2562657 | EPI2562658 | EPI2562659 | EPI2562660 | EPI2562661 | EPI2562662 | EPI2562663 |
| A/Kamigoto/920/2013 | EPI_ISL_17684817 | EPI2562648 | EPI2562649 | EPI2562650 | EPI2562651 | EPI2562652 | EPI2562653 | EPI2562654 | EPI2562655 |
| A/Kamigoto/878/2013 | EPI_ISL_17684804 | EPI2562640 | EPI2562641 | EPI2562642 | EPI2562643 | EPI2562644 | EPI2562645 | EPI2562646 | EPI2562647 |
| A/Kamigoto/817/2013 | EPI_ISL_17684803 | NA | NA | NA | EPI2562635 | EPI2562636 | EPI2562637 | EPI2562638 | EPI2562639 |
| A/Kamigoto/805/2013 | EPI_ISL_17684787 | NA | NA | NA | EPI2562630 | EPI2562631 | EPI2562632 | EPI2562633 | EPI2562634 |
| A/Kamigoto/722/2013 | EPI_ISL_17684786 | EPI2562622 | EPI2562623 | EPI2562624 | EPI2562625 | EPI2562626 | EPI2562627 | EPI2562628 | EPI2562629 |
| A/Kamigoto/707/2013 | EPI_ISL_17684785 | EPI2562606 | EPI2562607 | EPI2562608 | EPI2562609 | EPI2562610 | EPI2562612 | EPI2562614 | EPI2562616 |
| A/Kamigoto/667/2013 | EPI_ISL_17684676 | EPI2562597 | EPI2562598 | EPI2562599 | EPI2562600 | EPI2562601 | EPI2562602 | EPI2562603 | EPI2562604 |
| A/Kamigoto/666/2013 | EPI_ISL_17684609 | EPI2562584 | EPI2562585 | EPI2562587 | EPI2562588 | EPI2562589 | EPI2562590 | EPI2562591 | EPI2562593 |
| A/Kamigoto/665/2013 | EPI_ISL_17684607 | EPI2562575 | EPI2562577 | EPI2562578 | EPI2562579 | EPI2562580 | EPI2562581 | EPI2562582 | EPI2562583 |
| A/Kamigoto/635/2013 | EPI_ISL_17684606 | EPI2562567 | EPI2562568 | EPI2562569 | EPI2562570 | EPI2562571 | EPI2562572 | EPI2562573 | EPI2562574 |
| A/Kamigoto/624/2013 | EPI_ISL_17684520 | EPI2562559 | EPI2562560 | EPI2562561 | EPI2562562 | EPI2562563 | EPI2562564 | EPI2562565 | EPI2562566 |
| A/Kamigoto/618/2013 | EPI_ISL_17684519 | EPI2562551 | EPI2562552 | EPI2562553 | EPI2562554 | EPI2562555 | EPI2562556 | EPI2562557 | EPI2562558 |
| A/Kamigoto/617/2013 | EPI_ISL_17684518 | EPI2562543 | EPI2562544 | EPI2562545 | EPI2562546 | EPI2562547 | EPI2562548 | EPI2562549 | EPI2562550 |
| A/Kamigoto/616/2013 | EPI_ISL_17683953 | EPI2562535 | EPI2562536 | EPI2562537 | EPI2562538 | EPI2562539 | EPI2562540 | EPI2562541 | EPI2562542 |
| A/Kamigoto/615/2013 | EPI_ISL_17683857 | EPI2562527 | EPI2562528 | EPI2562529 | EPI2562530 | EPI2562531 | EPI2562532 | EPI2562533 | EPI2562534 |
| A/Kamigoto/614/2013 | EPI_ISL_17683792 | EPI2562519 | EPI2562520 | EPI2562521 | EPI2562522 | EPI2562523 | EPI2562524 | EPI2562525 | EPI2562526 |
| A/Kamigoto/612/2013 | EPI_ISL_17683791 | EPI2562511 | EPI2562512 | EPI2562513 | EPI2562514 | EPI2562515 | EPI2562516 | EPI2562517 | EPI2562518 |
| A/Kamigoto/603/2013 | EPI_ISL_17683790 | EPI2562503 | EPI2562504 | EPI2562505 | EPI2562506 | EPI2562507 | EPI2562508 | EPI2562509 | EPI2562510 |
| A/Kamigoto/600/2013 | EPI_ISL_17683728 | EPI2562495 | EPI2562496 | EPI2562497 | EPI2562498 | EPI2562499 | EPI2562500 | EPI2562501 | EPI2562502 |
| A/Kamigoto/590/2013 | EPI_ISL_17683727 | EPI2562487 | EPI2562488 | EPI2562489 | EPI2562490 | EPI2562491 | EPI2562492 | EPI2562493 | EPI2562494 |
| A/Kamigoto/561/2013 | EPI_ISL_17683726 | EPI2562479 | EPI2562480 | EPI2562481 | EPI2562482 | EPI2562483 | EPI2562484 | EPI2562485 | EPI2562486 |
| A/Kamigoto/480/2013 | EPI_ISL_17683724 | EPI2562463 | EPI2562464 | EPI2562465 | EPI2562466 | EPI2562467 | EPI2562468 | EPI2562469 | EPI2562470 |
| A/Kamigoto/475/2013 | EPI_ISL_17683723 | EPI2562455 | EPI2562456 | EPI2562457 | EPI2562458 | EPI2562459 | EPI2562460 | EPI2562461 | EPI2562462 |
| A/Kamigoto/375/2013 | EPI_ISL_17683722 | EPI2562447 | EPI2562448 | EPI2562449 | EPI2562450 | EPI2562451 | EPI2562452 | EPI2562453 | EPI2562454 |
| A/Kamigoto/369/2013 | EPI_ISL_17683294 | NA | NA | NA | EPI2562442 | NA | EPI2562444 | EPI2562445 | EPI2562446 |
| A/Kamigoto/310/2013 | EPI_ISL_17683293 | EPI2562434 | EPI2562435 | EPI2562436 | EPI2562437 | EPI2562438 | EPI2562439 | EPI2562440 | EPI2562441 |
| A/Kamigoto/296/2013 | EPI_ISL_17683292 | NA | NA | EPI2562428 | EPI2562429 | EPI2562430 | EPI2562431 | EPI2562432 | EPI2562433 |
| A/Kamigoto/274/2013 | EPI_ISL_17683291 | EPI2562420 | EPI2562421 | EPI2562422 | EPI2562423 | EPI2562424 | EPI2562425 | EPI2562426 | EPI2562427 |
| A/Kamigoto/273/2013 | EPI_ISL_17683048 | EPI2562412 | EPI2562413 | EPI2562414 | EPI2562415 | EPI2562416 | EPI2562417 | EPI2562418 | EPI2562419 |
| A/Kamigoto/220/2013 | EPI_ISL_17683047 | EPI2562404 | EPI2562405 | EPI2562406 | EPI2562407 | EPI2562408 | EPI2562409 | EPI2562410 | EPI2562411 |
| A/Kamigoto/216/2013 | EPI_ISL_17683046 | EPI2562396 | EPI2562397 | EPI2562398 | EPI2562399 | EPI2562400 | EPI2562401 | EPI2562402 | EPI2562403 |
| A/Kamigoto/208/2013 | EPI_ISL_17683045 | EPI2562388 | EPI2562389 | EPI2562390 | EPI2562391 | EPI2562392 | EPI2562393 | EPI2562394 | EPI2562395 |
| A/Kamigoto/1137/2012 | EPI_ISL_17296154 | EPI2477423 | EPI2477424 | EPI2477425 | EPI2477426 | EPI2477427 | EPI2477428 | EPI2477429 | EPI2477430 |
| A/Kamigoto/1209/2012 | EPI_ISL_17296116 | EPI2477415 | EPI2477416 | EPI2477417 | EPI2477418 | EPI2477419 | EPI2477420 | EPI2477421 | EPI2477422 |
| A/Kamigoto/1378/2012 | EPI_ISL_17296115 | EPI2477407 | EPI2477408 | EPI2477409 | EPI2477410 | EPI2477411 | EPI2477412 | EPI2477413 | EPI2477414 |
| A/Kamigoto/1430/2012 | EPI_ISL_17296114 | EPI2477399 | EPI2477400 | EPI2477401 | EPI2477402 | EPI2477403 | EPI2477404 | EPI2477405 | EPI2477406 |
| A/Kamigoto/1437/2012 | EPI_ISL_17296113 | EPI2477391 | EPI2477392 | EPI2477393 | EPI2477394 | EPI2477395 | EPI2477396 | EPI2477397 | EPI2477398 |
| A/Kamigoto/1439/2012 | EPI_ISL_17296112 | EPI2477383 | EPI2477384 | EPI2477385 | EPI2477386 | EPI2477387 | EPI2477388 | EPI2477389 | EPI2477390 |
| A/Kamigoto/1483/2012 | EPI_ISL_17296108 | EPI2477375 | EPI2477376 | EPI2477377 | EPI2477378 | EPI2477379 | EPI2477380 | EPI2477381 | EPI2477382 |
| A/Kamigoto/1490/2012 | EPI_ISL_17296105 | EPI2477367 | EPI2477368 | EPI2477369 | EPI2477370 | EPI2477371 | EPI2477372 | EPI2477373 | EPI2477374 |
| A/Kamigoto/1514/2012 | EPI_ISL_17296097 | EPI2477359 | EPI2477360 | EPI2477361 | EPI2477362 | EPI2477363 | EPI2477364 | EPI2477365 | EPI2477366 |
| A/Kamigoto/1530/2012 | EPI_ISL_17296096 | EPI2477351 | EPI2477352 | EPI2477353 | EPI2477354 | EPI2477355 | EPI2477356 | EPI2477357 | EPI2477358 |
| A/Kamigoto/894/2012 | EPI_ISL_17296073 | EPI2477343 | EPI2477344 | EPI2477345 | EPI2477346 | EPI2477347 | EPI2477348 | EPI2477349 | EPI2477350 |
| A/Kamigoto/887/2012 | EPI_ISL_17296061 | EPI2477335 | EPI2477336 | EPI2477337 | EPI2477338 | EPI2477339 | EPI2477340 | EPI2477341 | EPI2477342 |
| A/Kamigoto/886/2012 | EPI_ISL_17296025 | EPI2477327 | EPI2477328 | EPI2477329 | EPI2477330 | EPI2477331 | EPI2477332 | EPI2477333 | EPI2477334 |
| A/Kamigoto/871/2012 | EPI_ISL_17295949 | EPI2477319 | EPI2477320 | EPI2477321 | EPI2477322 | EPI2477323 | EPI2477324 | EPI2477325 | EPI2477326 |
| A/Kamigoto/805/2012 | EPI_ISL_17295903 | EPI2477311 | EPI2477312 | EPI2477313 | EPI2477314 | EPI2477315 | EPI2477316 | EPI2477317 | EPI2477318 |
| A/Kamigoto/796/2012 | EPI_ISL_17295902 | EPI2477303 | EPI2477304 | EPI2477305 | EPI2477306 | EPI2477307 | EPI2477308 | EPI2477309 | EPI2477310 |
| A/Kamigoto/784/2012 | EPI_ISL_17295671 | EPI2477295 | EPI2477296 | EPI2477297 | EPI2477298 | EPI2477299 | EPI2477300 | EPI2477301 | EPI2477302 |
| A/Kamigoto/772/2012 | EPI_ISL_17104742 | EPI2451325 | EPI2451326 | EPI2451327 | EPI2451328 | EPI2451329 | EPI2451330 | EPI2451331 | EPI2451332 |
| A/Kamigoto/771/2012 | EPI_ISL_17104741 | EPI2451317 | EPI2451318 | EPI2451319 | EPI2451320 | EPI2451321 | EPI2451322 | EPI2451323 | EPI2451324 |
| A/Kamigoto/765/2012 | EPI_ISL_17104740 | EPI2451309 | EPI2451310 | EPI2451311 | EPI2451312 | EPI2451313 | EPI2451314 | EPI2451315 | EPI2451316 |
| A/Kamigoto/763/2012 | EPI_ISL_17104739 | EPI2451301 | EPI2451302 | EPI2451303 | EPI2451304 | EPI2451305 | EPI2451306 | EPI2451307 | EPI2451308 |
| A/Kamigoto/762/2012 | EPI_ISL_17104738 | EPI2451293 | EPI2451294 | EPI2451295 | EPI2451296 | EPI2451297 | EPI2451298 | EPI2451299 | EPI2451300 |
| A/Kamigoto/757/2012 | EPI_ISL_17104737 | EPI2451285 | EPI2451286 | EPI2451287 | EPI2451288 | EPI2451289 | EPI2451290 | EPI2451291 | EPI2451292 |
| A/Kamigoto/752/2012 | EPI_ISL_17104735 | EPI2451277 | EPI2451278 | EPI2451279 | EPI2451280 | EPI2451281 | EPI2451282 | EPI2451283 | EPI2451284 |
| A/Kamigoto/1460/2012 | EPI_ISL_17103548 | EPI2449375 | EPI2449376 | EPI2449377 | EPI2449378 | EPI2449379 | EPI2449380 | EPI2449381 | EPI2449382 |
| A/Kamigoto/749/2012 | EPI_ISL_17103148 | EPI2449359 | EPI2449360 | EPI2449361 | EPI2449362 | EPI2449363 | EPI2449364 | EPI2449365 | EPI2449366 |
| A/Kamigoto/729/1012 | EPI_ISL_17103065 | EPI2449342 | EPI2449344 | EPI2449345 | EPI2449346 | EPI2449347 | EPI2449348 | EPI2449349 | EPI2449350 |
| A/Kamigoto/175/2013 | EPI_ISL_17092239 | EPI2440003 | EPI2440052 | EPI2440085 | EPI2440098 | EPI2440099 | EPI2440100 | EPI2440101 | EPI2440102 |
| A/Kamigoto/1075/1012 | EPI_ISL_17103062 | EPI2449319 | EPI2449320 | EPI2449321 | EPI2449322 | EPI2449323 | EPI2449324 | EPI2449325 | EPI2449326 |

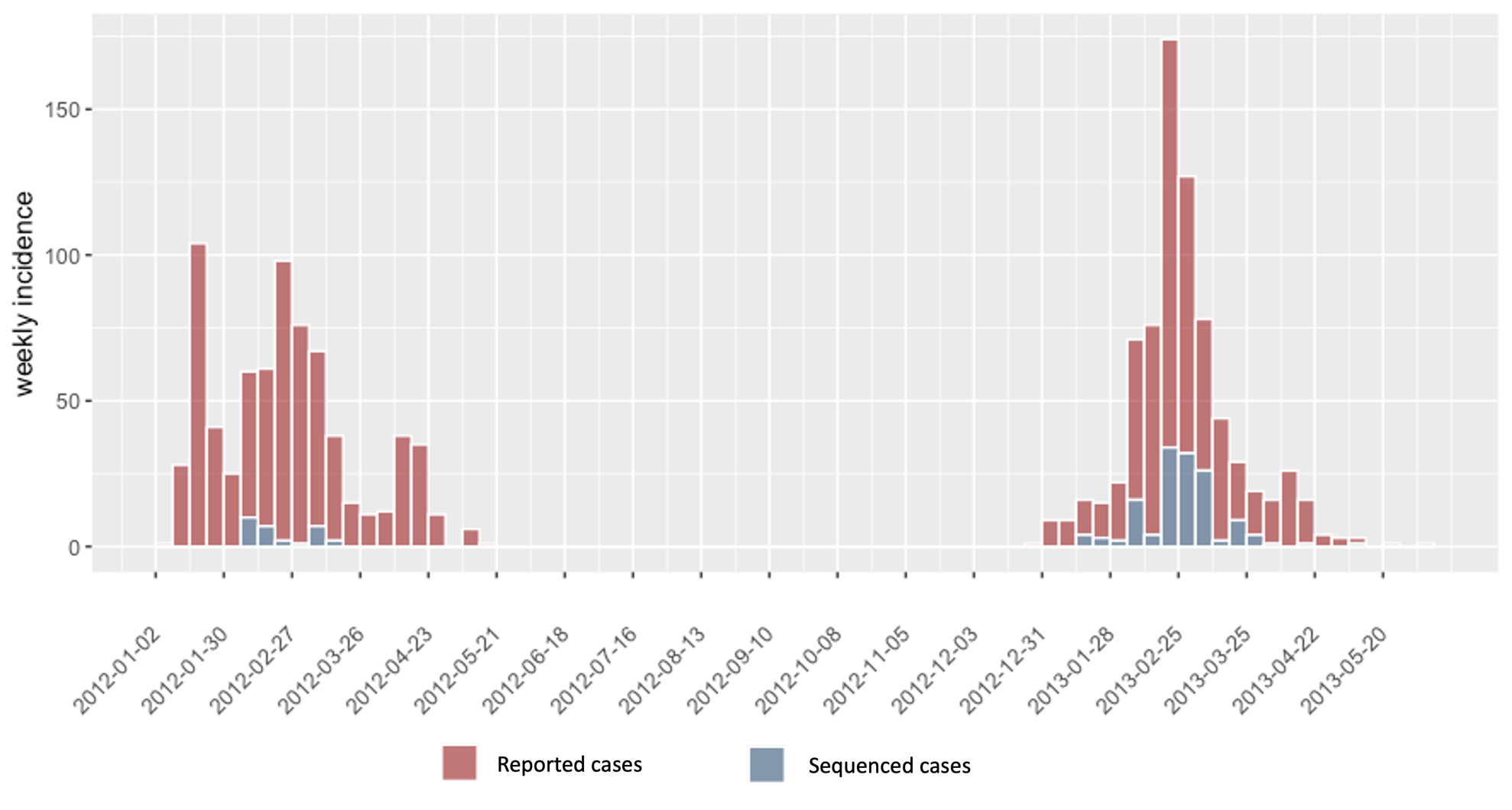

**Figure S-1:** Epidemic curve of the daily number of rapid influenza diagnostic test positive cases reported by the Hospital influenza surveillance system (red), and whole genome sequenced (WGS) cases (blue) during the 2011/12 and 2012/2013 epidemic seasons.

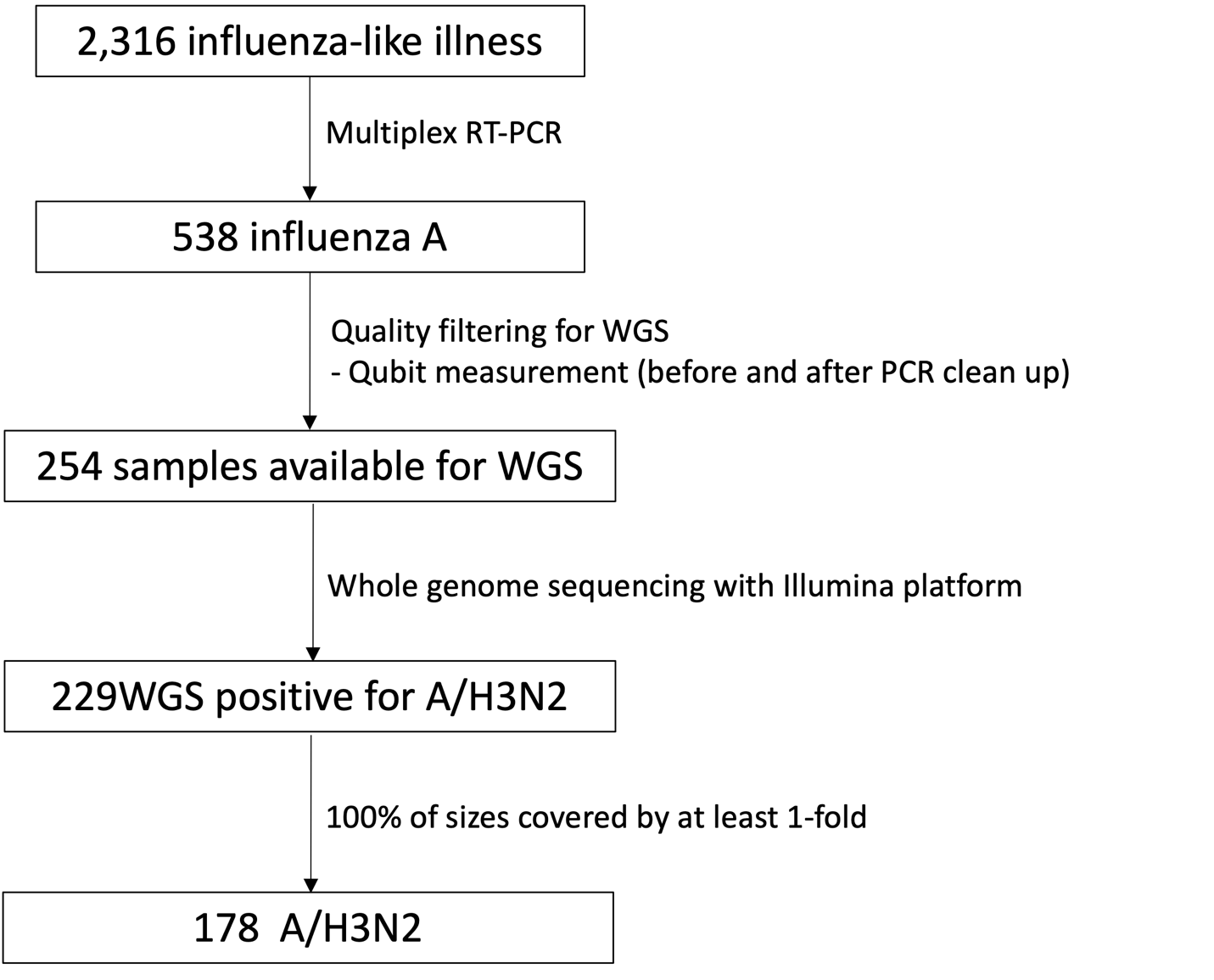

**FIGURE S-2:** Flowchart showing from samples collection to whole genome sequencing (WGS)

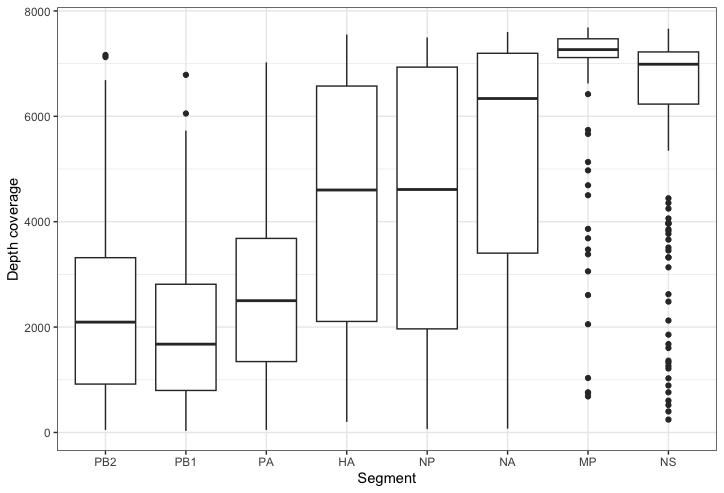

**FIGURE S-3:** Boxplots illustrating median read depth across each segment for A/H3N2 virus Boxes extend to the 1^st^ and 3^rd^ quartile.

**FIGURE S-4.** The maximum likelihood phylogenetic tree includes 168 WGS collected during the 2011/12 (red) and 2012/13 (purple) influenza seasons alongside the WGS in Japan, that were available in the GISAID (blue), and the vaccine strains (green). For each sequence the date of the sample collection is mentioned (yyyy-mm-dd). WGS for 2012/2013 in Japan were not available during the study period.

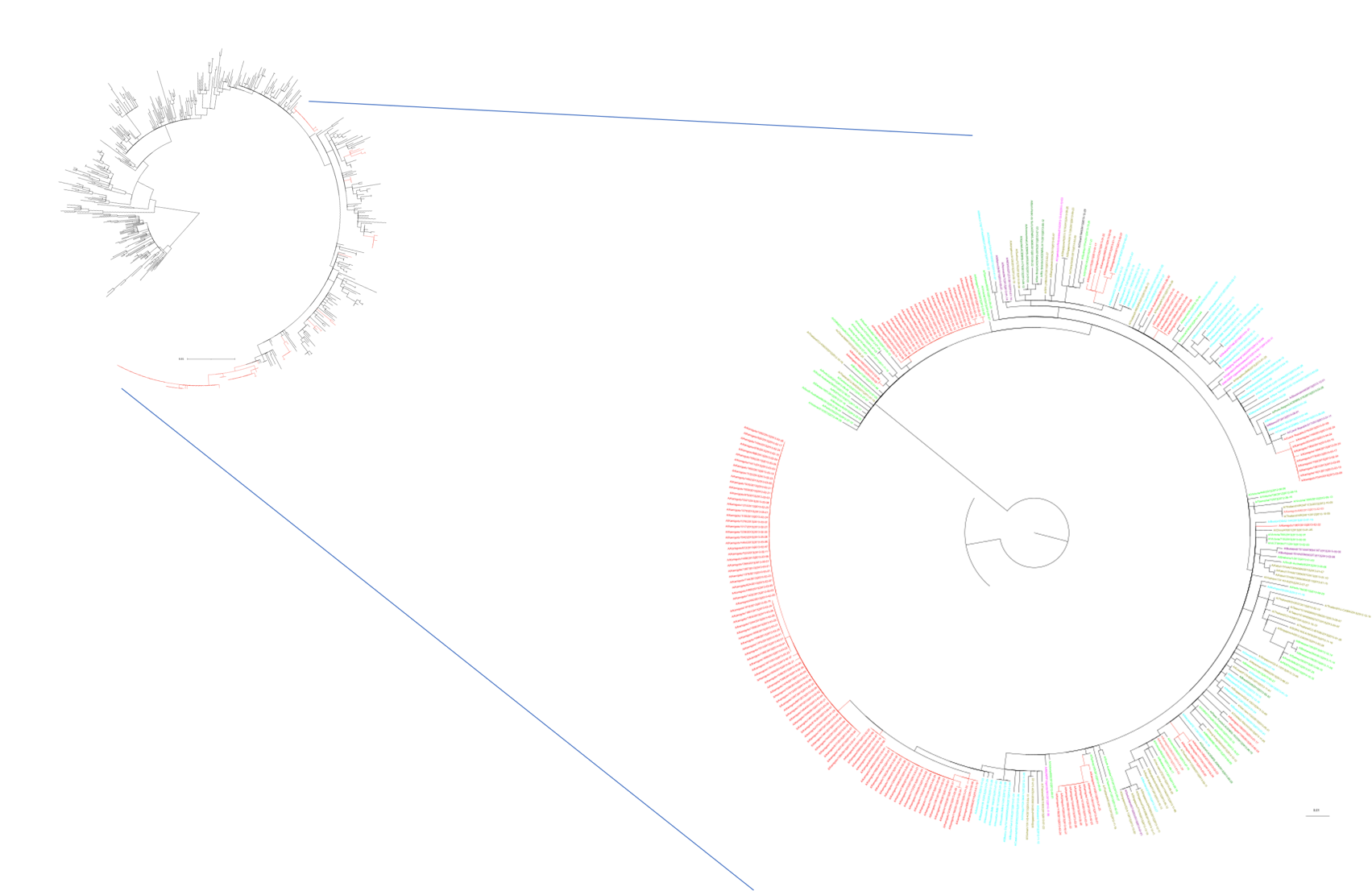

**Figure S-5:** **Maximum-likelihood phylogenetic trees of PB2 segments of influenza A/H3N2 viruses circulating in Kamigoto and comparing sequences from strains isolated in Japan and other parts of the world from GISAID collected between 2011 and 2013.**

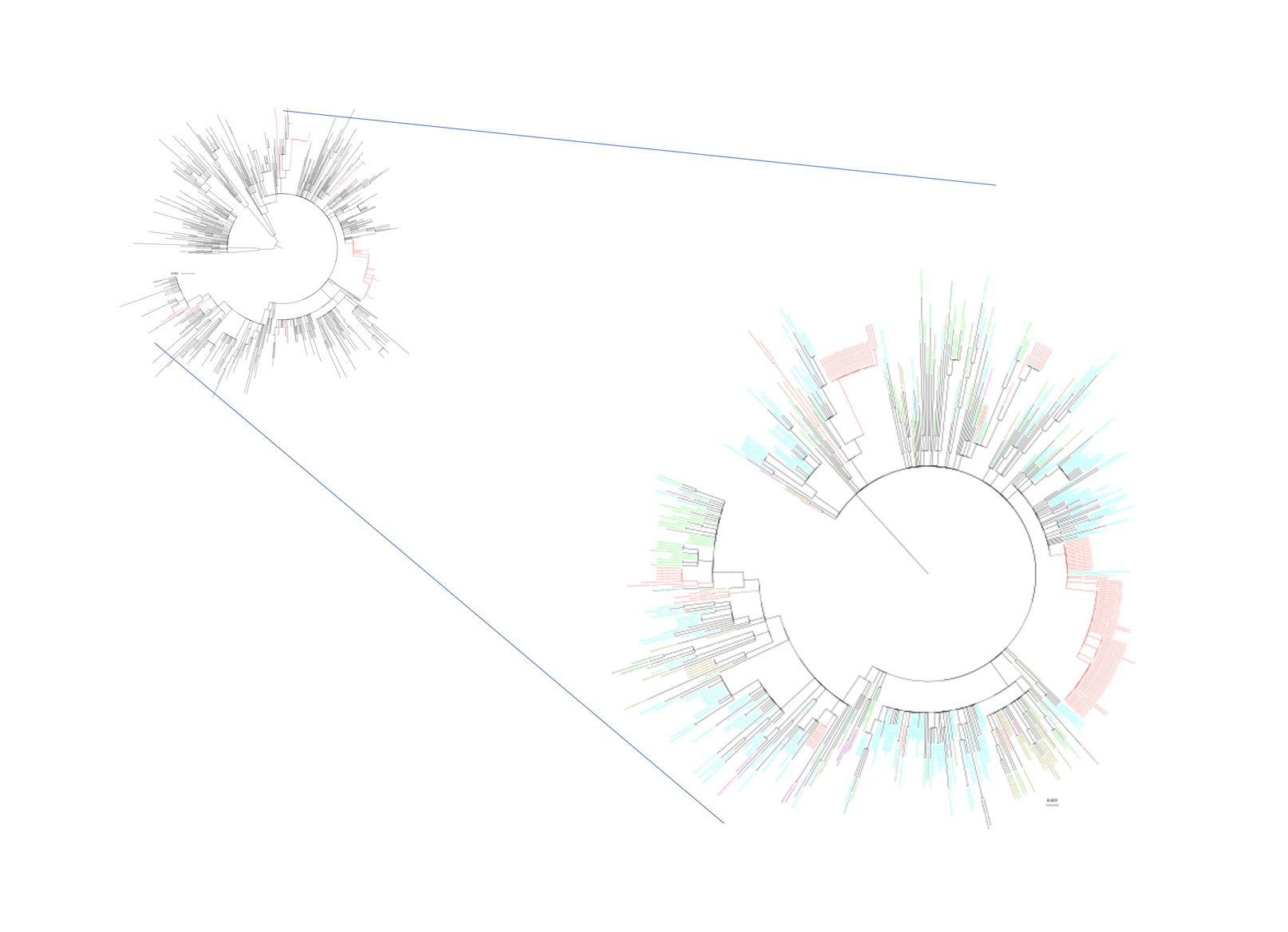

**Figure S-6:** **Maximum-likelihood phylogenetic trees of PB1 segments of influenza A/H3N2 viruses circulating in Kamigoto and comparing sequences from strains isolated in Japan and other parts of the world from GISAID collected between 2011 and 2013.**

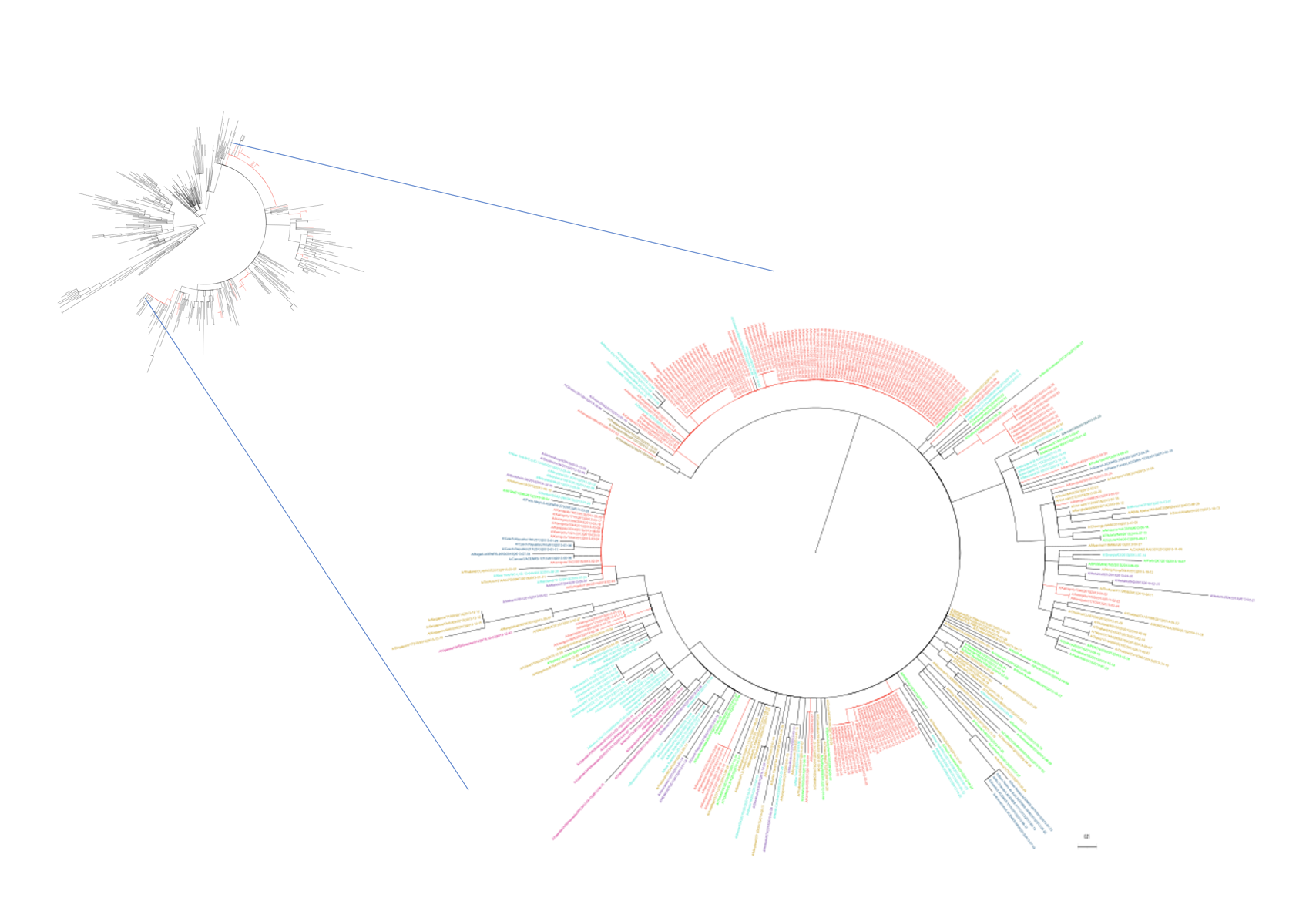

**Figure S-7:** **Maximum-likelihood phylogenetic trees of PA segments of influenza A/H3N2 viruses circulating in Kamigoto and comparing sequences from strains isolated in Japan and other parts of the world from GISAID collected between 2011 and 2013.**

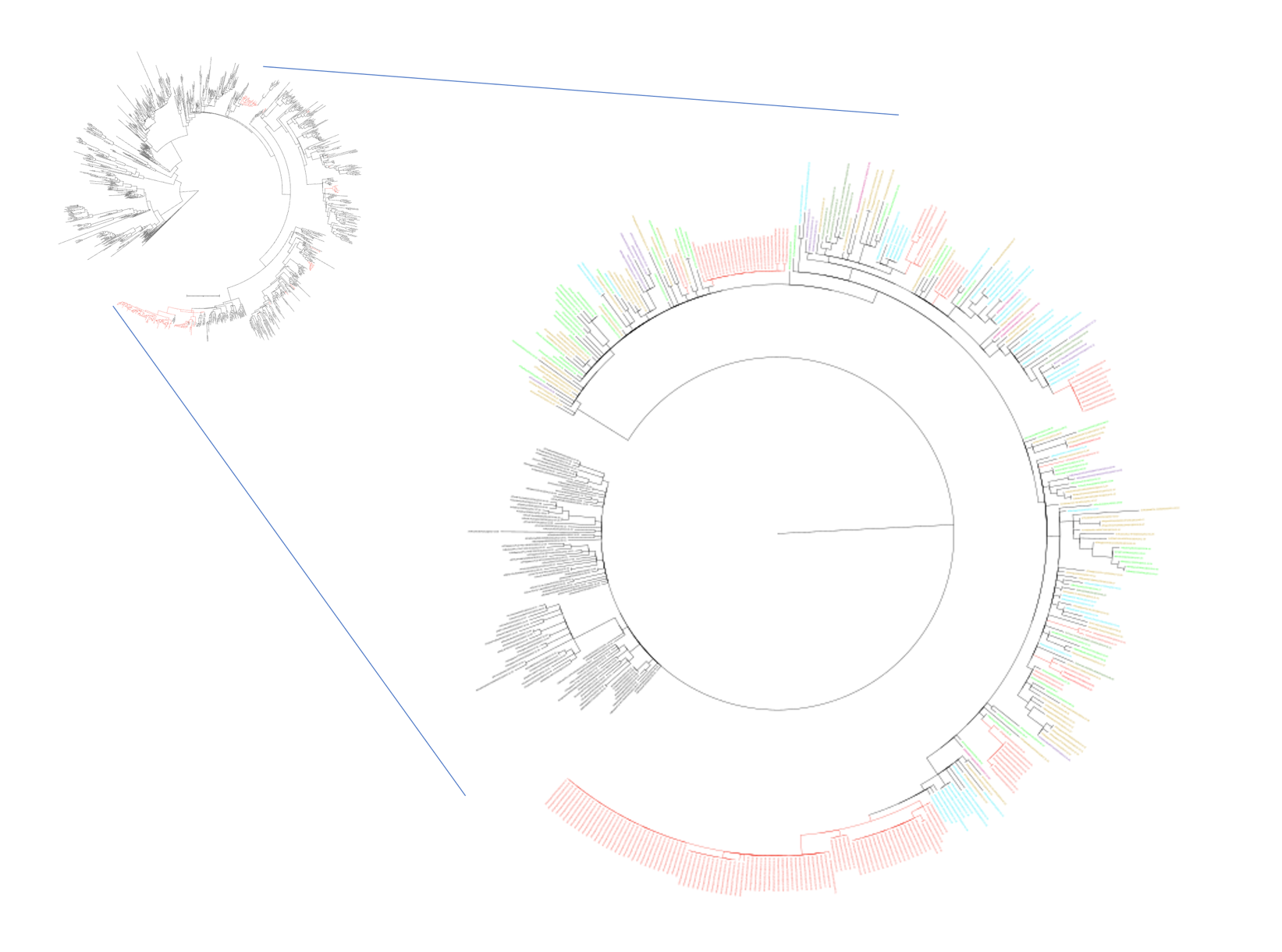

**Figure S-8:** **Maximum-likelihood phylogenetic trees of HA segments of influenza A/H3N2 viruses circulating in Kamigoto and comparing sequences from strains isolated in Japan and other parts of the world from GISAID collected between 2011 and 2013.**

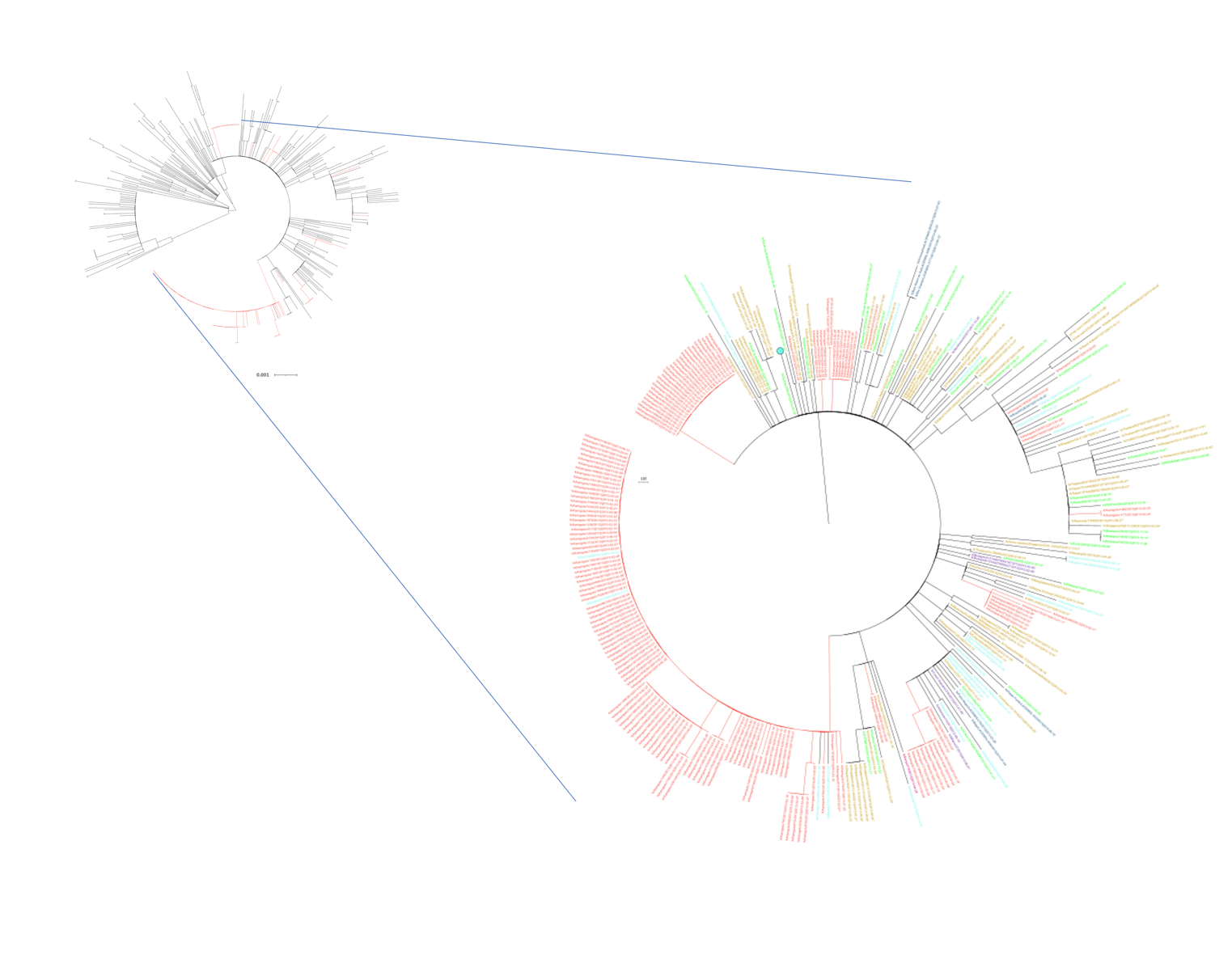

**Figure S-9:** **Maximum-likelihood phylogenetic trees of NP segments of influenza A/H3N2 viruses circulating in Kamigoto and comparing sequences from strains isolated in Japan and other parts of the world from GISAID collected between 2011 and 2013.**

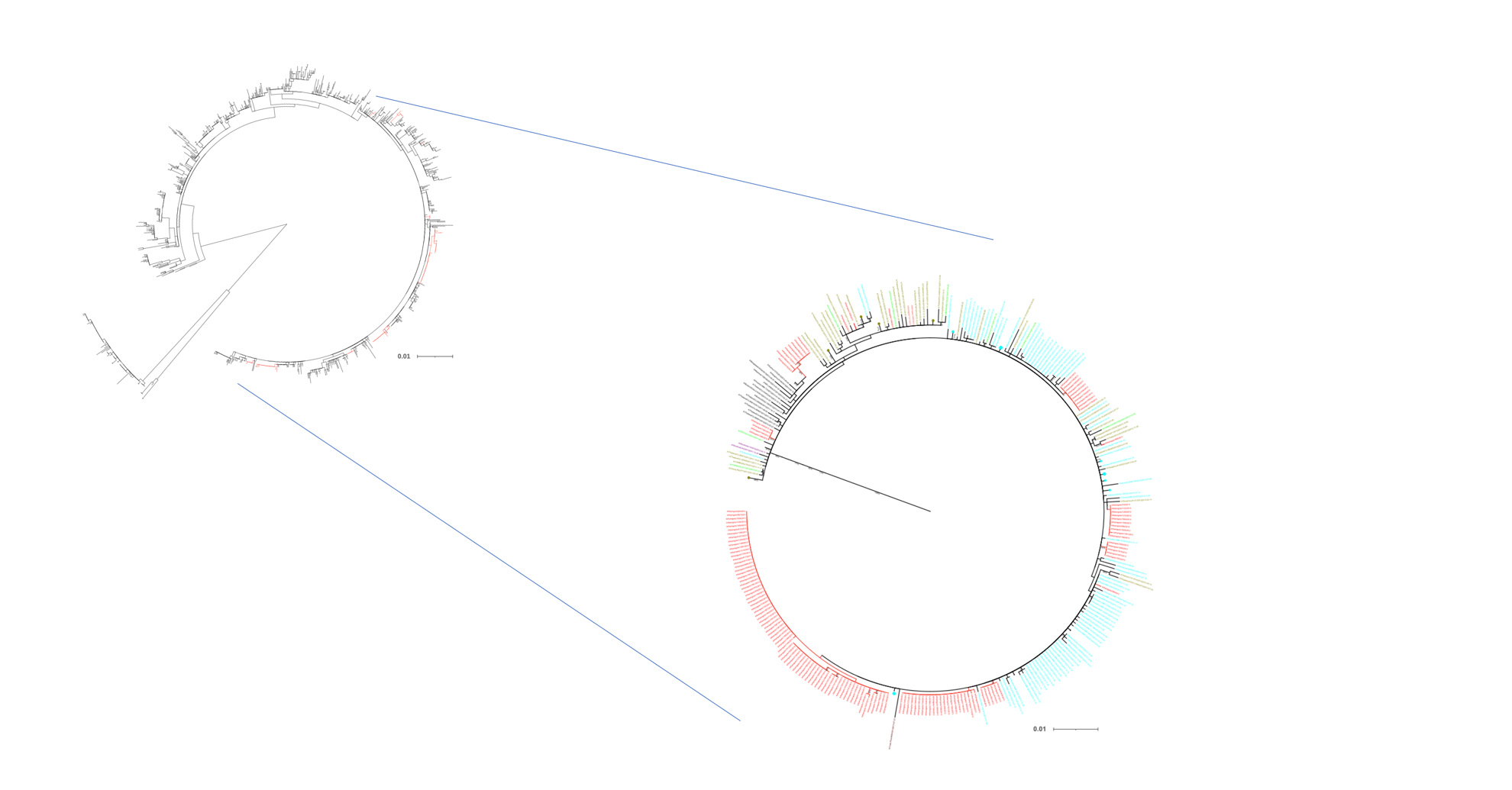

**Figure S-10:** **Maximum-likelihood phylogenetic trees of NA segments of influenza A/H3N2 viruses circulating in Kamigoto and comparing sequences from strains isolated in Japan and other parts of the world from GISAID collected between 2011 and 2013.**

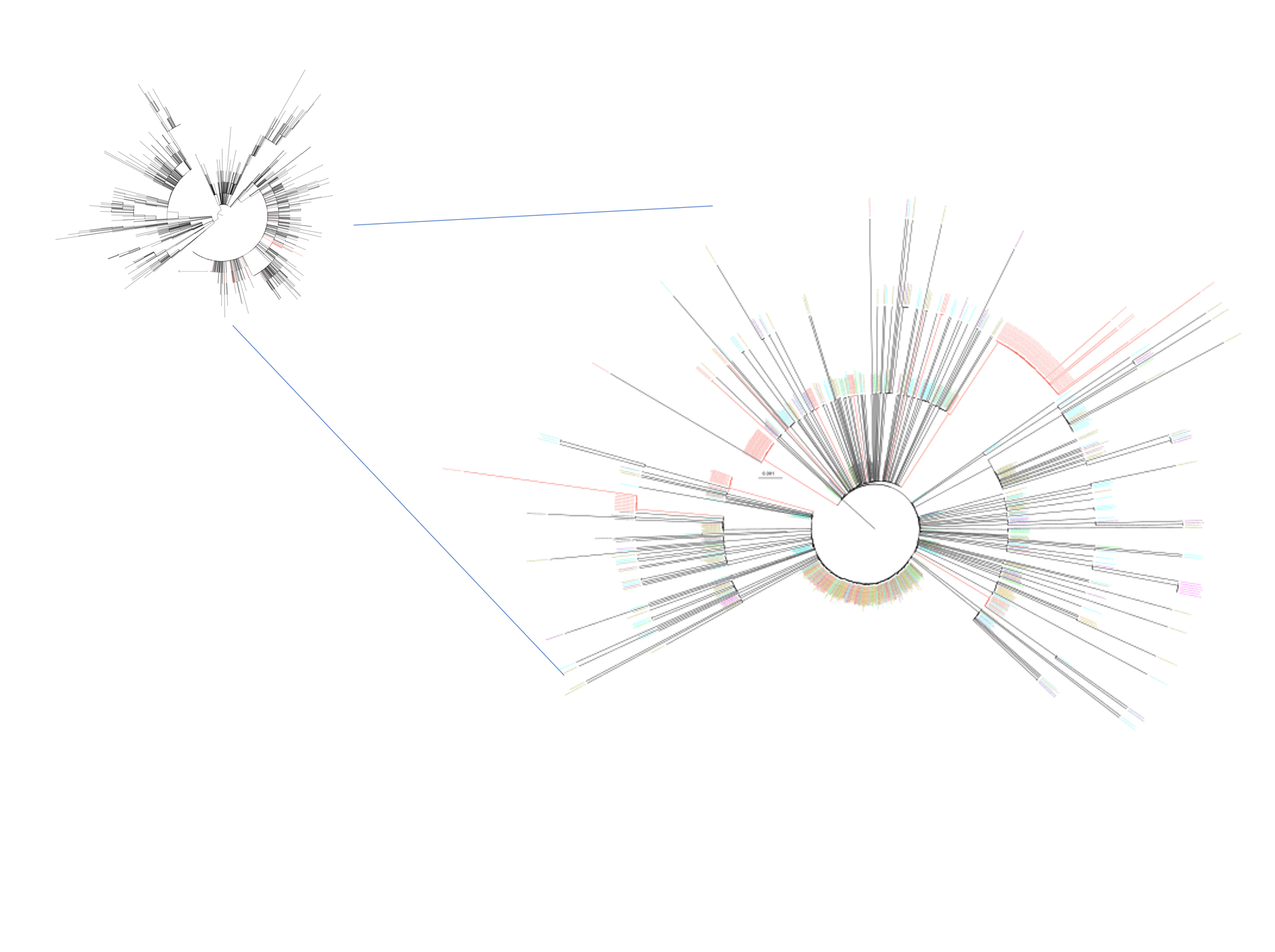

**Figure S-11:** **Maximum-likelihood phylogenetic trees of MP segments of influenza A/H3N2 viruses circulating in Kamigoto and comparing sequences from strains isolated in Japan and other parts of the world from GISAID collected between 2011 and 2013.**

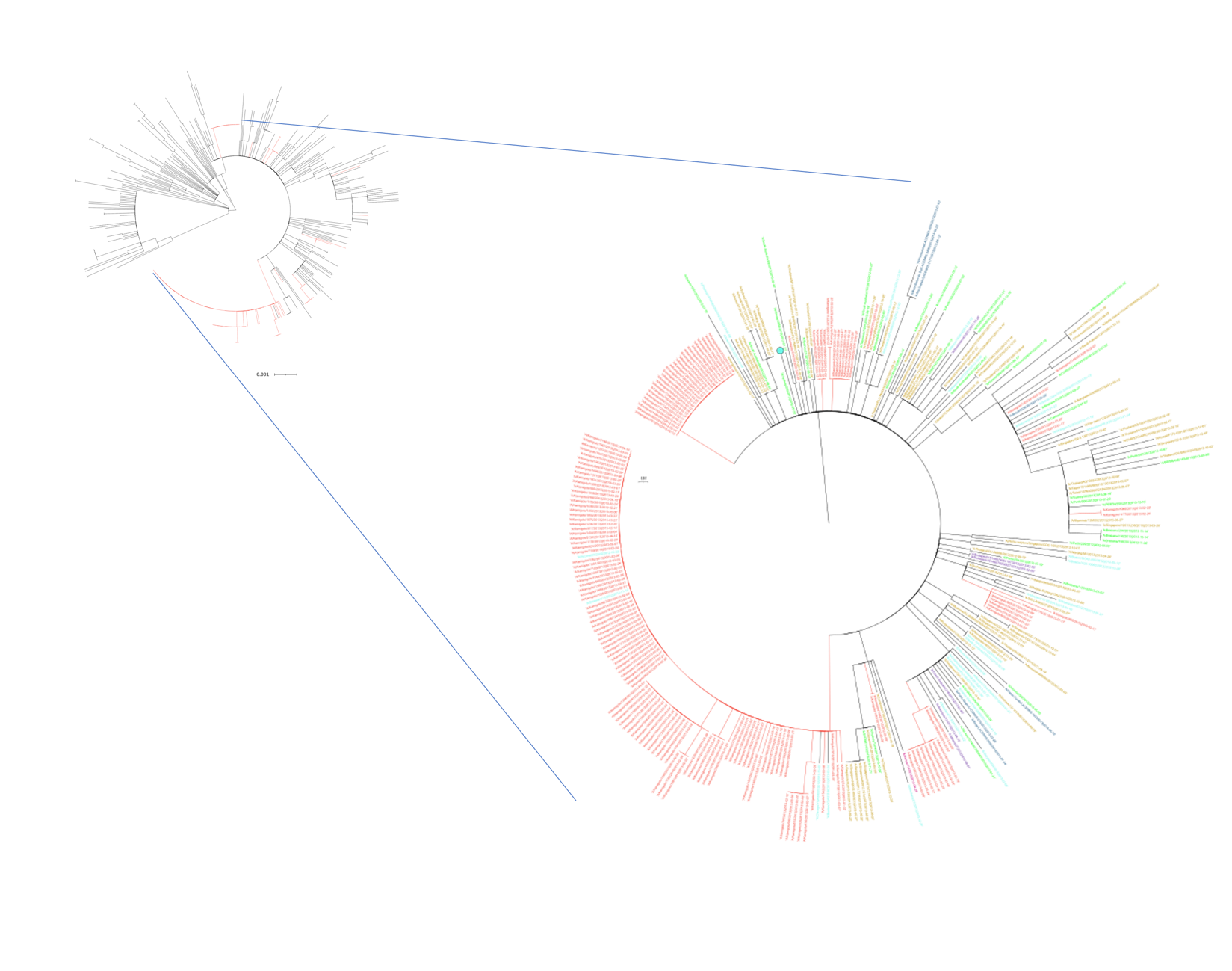

**Figure S-12:** **Maximum-likelihood phylogenetic trees of NS segments of influenza A/H3N2 viruses circulating in Kamigoto and comparing sequences from strains isolated in Japan and other parts of the world from GISAID collected between 2011 and 2013.**

The figure is in tiff file as separate document

**FIGURE S-13.** Time resolved phylogenetic tree of HA segments of Kamigoto island, Japan and global sequences (GISAID) as of 1^st^ December 2022 submission date.

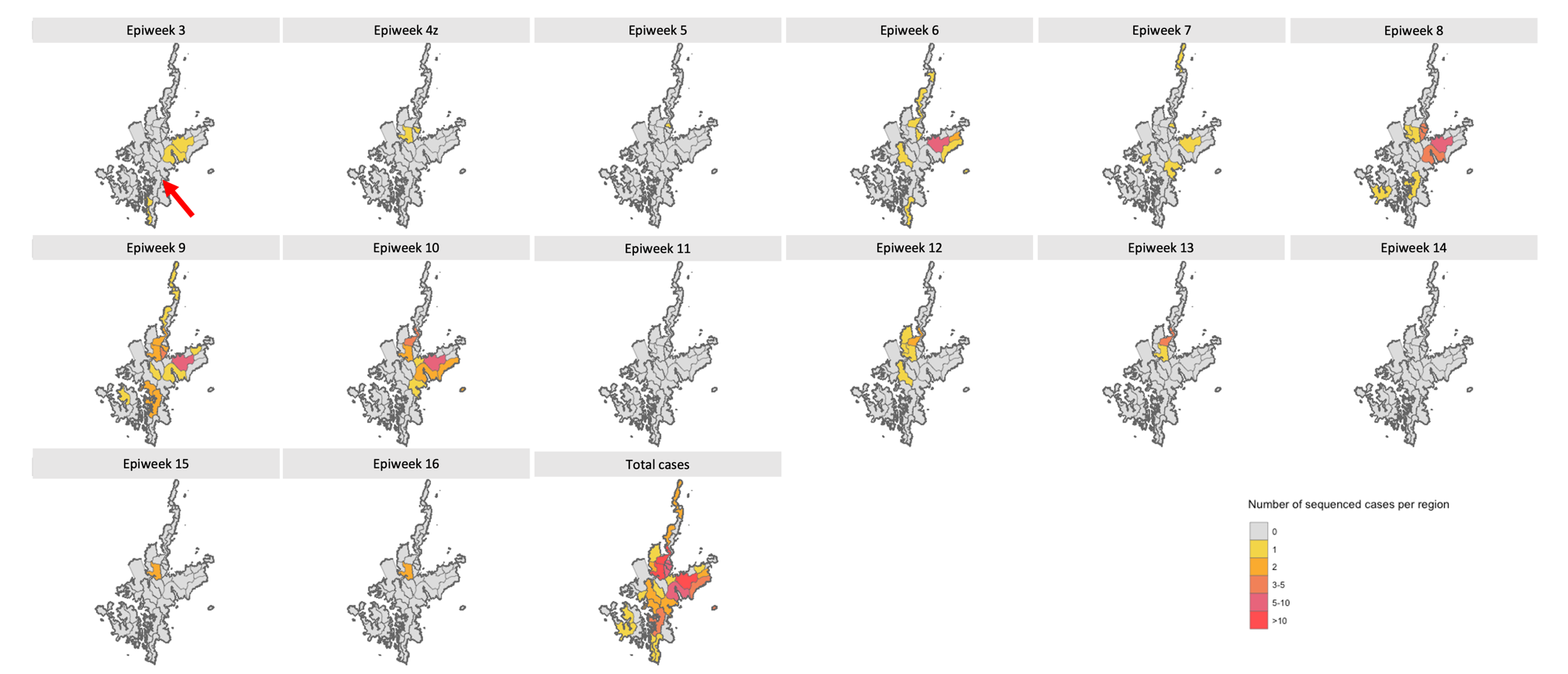

**Figure S-14:** Temporal and spatial distribution of the sequenced cases of cluster 5 (5A and 5B) (weekly)

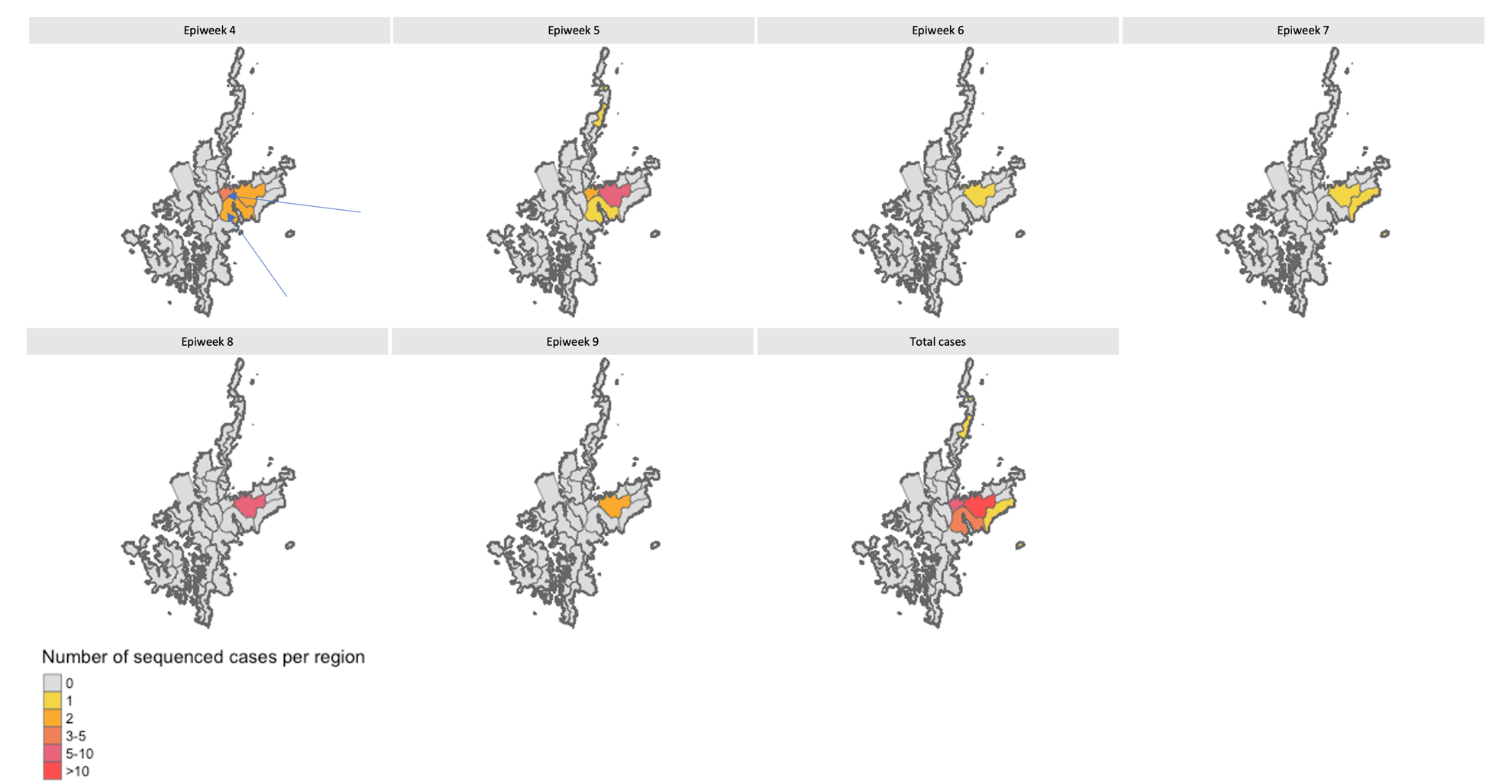

**Figure S-15:** Temporal and spatial distribution of the sequenced cases of cluster 1 (weekly)
